# Bi-allelic variants in the aminopeptidase XPNPEP3 cause mitochondrial disease with pediatric cardiomyopathy

**DOI:** 10.1101/2025.01.11.25320052

**Authors:** Claudine W.B. Ruijmbeek, Sjoerd Ruizenaar, Herma C. van der Linde, Edgar E. Nollet, Wouter A.S. Doff, Victoria C.S. Bogaard, Marlène de Pee, Federico Ferraro, Richard J. Rodenburg, Henk S. Schipper, Alexander Hirsch, Marjon A. van Slegtenhorst, Jan H. von der Thüsen, Jeroen A.A. Demmers, Wilfred F.J. van IJcken, Tjakko J. van Ham, Judith M. A. Verhagen

## Abstract

Bi-allelic variants in *XPNPEP3* are known to cause nephronophthisis-like nephropathy-1. However, recent findings indicate XPNPEP3 deficiency causes a broader phenotypic spectrum encompassing extra-renal manifestations. XPNPEP3 is suggested to be the human ortholog of ICP55, a mitochondrial protein involved in the processing and maturation of mitochondrial proteins in plants and yeast.

Here, we present a family with two children affected by a homozygous splice variant in *XPNPEP3*. Both children exhibited early-onset renal insufficiency and progressive mixed hypertrophic and dilated cardiomyopathy, requiring cardiac transplantation during childhood. RNA and protein analysis of patient fibroblasts and cardiac tissue revealed loss of XPNPEP3 expression due to the splice variant. Assessment of explanted cardiac tissue confirmed mitochondrial dysfunction, indicated by decreased cytochrome c oxidase activity and changes in mitochondrial morphology.

Although the deficiency of Xpnpep3 in zebrafish did not result in noticeable phenotypic abnormalities during early larval stages, transcriptomic and proteomic analyses revealed mitochondrial alterations. Notably, proteomic profiling identified decreased abundance of mitochondrial proteins in *xpnpep3^Δ7/Δ7^* mutants as compared to wild-types, including those identified in our study as putative substrates of Xpnpep3. We hypothesize that impaired processing of these proteins plays a critical role in the early developmental molecular changes observed, potentially predisposing to disease and also underlies the clinical manifestations in affected children.

In summary, our study underscores that loss of XPNPEP3 causes a mitochondrial disorder with varied phenotypes, including cardiomyopathy, through a molecular mechanism likely involving abnormal processing and stabilization of mitochondrial precursor proteins. Adding *XPNPEP3* to cardiac and mitochondrial disease gene panels is essential for the accurate diagnosis and management of potential cardiac complications.

## Introduction

Mitochondrial diseases are a diverse group of disorders arising from variants in nuclear and/or mitochondrial DNA (mtDNA). The mitochondrial proteome consists of approximately 1100 to 1500 proteins and about 400 of these are disease-related [1–3]. Most mitochondrial diseases are caused by pathogenic variants in the genes encoding subunits of the oxidative phosphorylation system (OXPHOS) or in genes encoding proteins that are functionally connected with the OXPHOS, resulting in defective adenosine triphosphate (ATP) energy production [4]. OXPHOS is made of four multi-subunit enzyme complexes (also referred to as complex I to IV) that form the respiratory chain, two mobile electron carriers, and ATP synthase (known as complex V) [5]. Diagnosing individuals with mitochondrial disease poses a significant challenge for clinicians because symptoms can vary and are often nonspecific. The clinical presentation is highly heterogeneous, ranging from isolated organ involvement to simultaneous impacts on multiple systems, and particularly affects organs with high energy demands, such as the brain, and skeletal and cardiac muscle. Symptoms also vary in severity and age of onset; progression may be rapid and severe in some cases, while stable and well-managed in others [6]. Last, phenotypic heterogeneity among individuals sharing the same genetic variant further complicates the diagnosis and management of mitochondrial disorders [7–9].

Cardiac complications, such as cardiomyopathy and cardiac conduction defects, are major contributors to mortality in individuals affected by mitochondrial disease, especially in the pediatric population [10–12]. Studies suggest that 20–40% of pediatric patients with mitochondrial disease develop cardiomyopathy, thereby worsening their prognosis [13, 14]. Cardiomyopathies can be categorized based on their morphological and functional characteristics. The predominant types include hypertrophic cardiomyopathy (HCM) and dilated cardiomyopathy (DCM) [15]. Notably, over 40% of identified disease genes associated with mitochondrial disease have been shown to cause a form of cardiomyopathy [16–19]. These genes code for proteins that are involved in mitochondrial structure and function (e.g. OXPHOS, mitochondrial protein import, homeostasis and quality control), but also in mtDNA maintenance, expression, and translation.

In the present study, family-based exome sequencing in two siblings with renal insufficiency and rapidly progressive cardiomyopathy revealed a novel homozygous variant (NM_022098.4 c.1357G>A) in the *XPNPEP3* gene, which we showed to cause abnormal splicing, leading to an abnormal transcript and loss of XPNPEP3 protein function. Bi-allelic variants in *XPNPEP3* are associated with nephronophthisis-like nephropathy-1 (NPHPL1, MIM 613159). O’Toole et al. proposed that the pathogenesis of this condition is driven by defects in ciliary proteins as a result of XPNPEP3 dysfunction [20]. Nevertheless, recent studies indicate a broader phenotypic spectrum that includes extra-renal manifestations, including neuromuscular symptoms [20–24]. Cardiomyopathy has been observed in three affected children from two of these reported families [20, 24]. Two siblings carried a homozygous loss-of-function (LoF) variant, whereas the child from the other family had a homozygous splice variant. The child from the latter family died from heart failure [24].

The *XPNPEP3* gene encodes X-prolyl aminopeptidase and is proposed to be the human ortholog of yeast intermediate-cleavage peptidase 55 (ICP55) [25]. Studies in yeast and plants have characterized the role of ICP55 in processing nuclear-encoded mitochondrial proteins [25–31]. Nuclear-encoded mitochondrial proteins are synthesized on cytosolic ribosomes and subsequently transported into the mitochondria. These precursor proteins contain a N-terminal mitochondrial targeting signal (MTS), which serve as a guiding signal directing their import into the mitochondrial matrix. Upon entry into the mitochondrial matrix, these precursor proteins undergo cleavage by specialized proteases. The mitochondrial processing peptidase (MPP) cleaves off a major part of the MTS [1, 2]. Some of the precursor proteins require an additional processing step, carried out by either mitochondrial intermediate peptidase (OCT1 in yeast, MIPEP in humans), cleaving of octapeptides, or ICP55, cleaving off a single amino acid, leading to stable mature proteins [32–34]. While XPNPEP3 has been shown to localize within mitochondria in rat renal cells, and to possess structural and in vitro enzymatic substrate similarities to ICP55, there is currently limited evidence supporting this function in humans [20, 25]. To date, it remains unclear whether XPNPEP3 has a similar function to ICP55 in humans and if its dysfunction leads to cardiomyopathy.

Here, we report a third family with *XPNPEP3*-related cardiomyopathy. Two siblings developed end-stage heart failure requiring cardiac transplantation. Molecular analysis identified a predicted homozygous missense variant, which we demonstrated to cause abnormal splicing resulting in loss of XPNPEP3 protein expression. Using cardiac tissue from both affected siblings, consistent mitochondrial dysfunction was detected, evidenced by reduced complex IV (cytochrome c oxidase) activity and aberrant mitochondrial morphology. Moreover, our Xpnpep3-deficient zebrafish model showed mitochondrial alterations at both transcriptomic and proteomic levels during early development. Proteomic analyses in these zebrafish showed a reduced abundance of mitochondrial proteins that we identified as potential substrates of Xpnpep3. We hypothesize that their improper processing may explain their lower abundance and other molecular changes identified in mitochondrial and metabolic pathways. These findings should be validated using advanced mass-spectrometry techniques and further studies on patient-derived tissues. Overall, our study underscores the critical need to include *XPNPEP3* in cardiac and mitochondrial disease gene panels. Given the potentially life-threatening cardiac complications, we recommend regular cardiac assessment in patients with bi-allelic loss-of-function *XPNPEP3* variants. Identifying more patients with *XPNPEP3* variants is essential to gain a comprehensive understanding of this mitochondrial disorder.

## Results

### Clinical presentation

The proband (individual III:2; **Figure 1A**), the second child of consanguineous parents, was diagnosed with B-thalassemia, microcytic anemia, hyperparathyroidism, and renal insufficiency in childhood. In early adolescence (age range: 11–15 years), the proband developed acute heart failure. High-sensitivity cardiac troponin T (hs-cTnT) and N-terminal proB-type natriuretic peptide (NT-proBNP) levels were elevated, indicative of myocardial stress and heart failure **(Supplemental Figure 1, A and B)**. The electrocardiogram displayed sinus rhythm at 100 beats/min, a wide p-wave indicative of left atrial enlargement, a normal heart axis, normal atrioventricular conduction, normal QRS voltages in the precordial leads, and nonspecific repolarization abnormalities. Transthoracic echocardiography showed left ventricular (LV) dilatation (LV internal diameter end-diastole (LVIDd) 61 mm, z-score 6.4 [35]), with severely reduced systolic function (LV ejection fraction (LVEF) 17%), and severe left atrial enlargement. The proband was admitted to the intensive care unit for pharmacological and mechanical support. Due to persistent pulmonary edema, a left ventricular assist device was required, followed by uncomplicated cardiac transplantation (HTx). Post-HTx, the proband developed an adequate graft function and there were no signs of rejection on standard immunosuppression. Three years after transplantation however, the first signs of cardiac allograft vasculopathy appeared, and percutaneous stenting of the left anterior descending coronary artery was performed. Moreover, the kidney function is slowly declining, and renal replacement therapy and/or kidney transplantation may be needed in the future, despite a tacrolimus-sparing immunosuppressive regimen.

**Figure 1.**
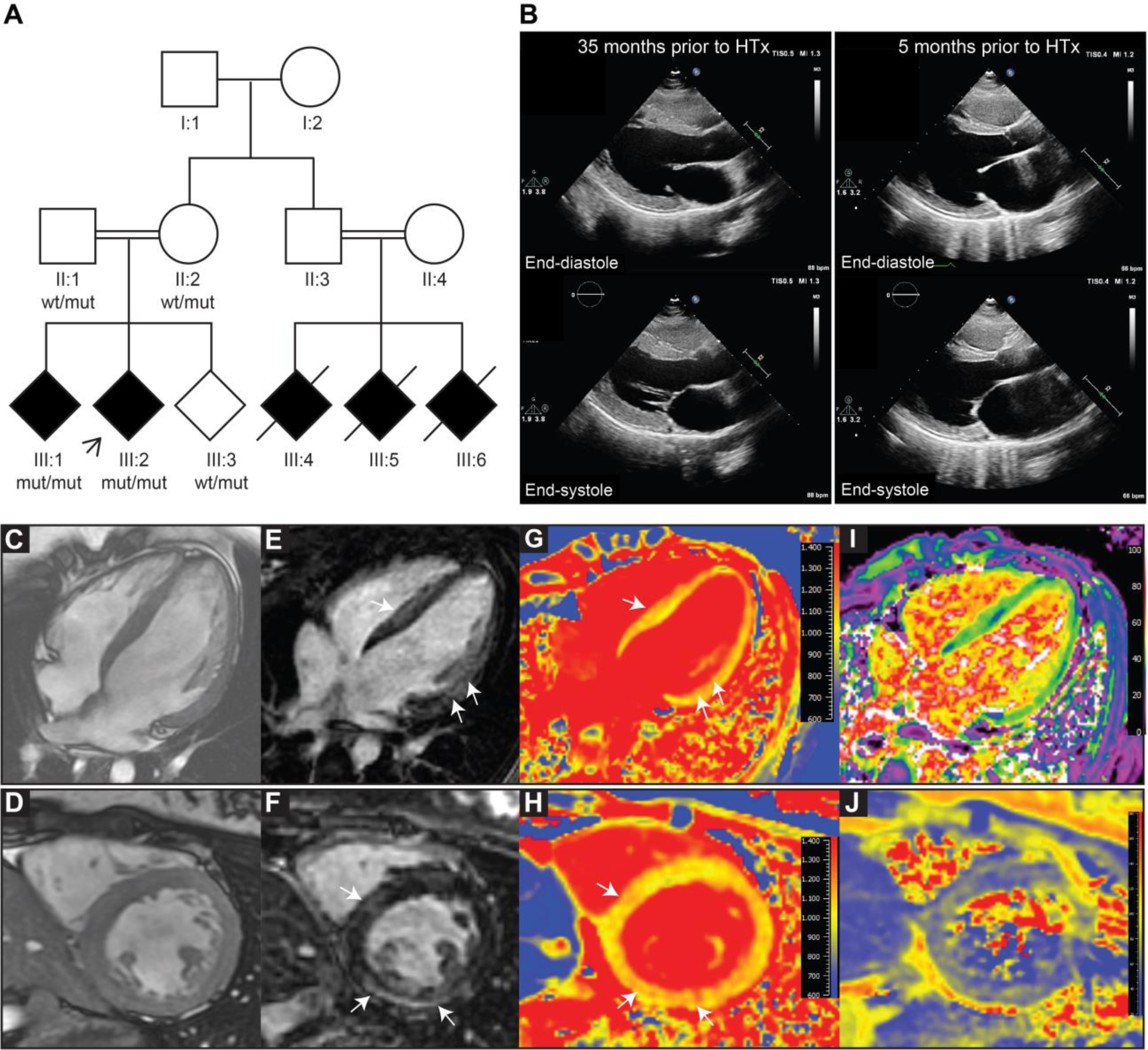
Clinical presentation. (A) Family pedigree of affected individuals. The arrowhead indicates the proband. (B) Cardiac ultrasound two-dimensional, parasternal long-axis transthoracic echocardiographic images of individual III:1, assessed 35 months (left) and 5 months (right) prior to cardiac transplantation. (C-J) Cardiovascular magnetic resonance (CMR) of individual III:1, with end-diastolic balanced steady-state free precession cine images in the four-chamber (C) and mid-ventricular short-axis views (D). (E, F) Corresponding late gadolinium enhancement (LGE) images. The white arrows highlight LGE in the mid-wall and epicardial regions of the septum and lateral wall. (G,H) Corresponding native T1 maps with increased T1 values (white arrows). (I) Extracellular volume (ECV) map in the 4 chamber view with diffuse increased ECV. (J) T2 map in mid-ventricular shot axis view with slightly elevated T2 values.

At the time of the proband’s HTx, first-degree relatives underwent comprehensive cardiac screening revealing no abnormalities in the parents and younger sibling, except for mild left atrial dilatation in one of the parents. Echocardiography performed in the older sibling (individual III:1; **Figure 1A**) during adolescence revealed moderate LV dysfunction (LVEF 44%) and normal LV dimensions (LVIDd: 48 mm, z-score -0.14) (**Figure 1B**, left panels). Cardiovascular magnetic resonance (CMR) revealed concentric LV hypertrophy, with an interventricular septum thickness of 13 mm, global systolic impairment (LVEF 41%; normal 54% [36]), and mild LV dilatation (LV end diastolic volume 93 ml/m2; upper limit normal 85 ml/m2 [36]) **(Figure 1, C and D)**. Late gadolinium enhancement (LGE) imaging further showed mid-wall and epicardial LGE in the septal, inferior and infero-lateral regions **(Figure 1, E and F)**. Parametric mapping showed increased native T1 times, elevated extracellular volume (ECV), and slightly elevated T2 times indicative of myocardial edema and fibrosis **(Figure 1, G-J)**. In the following years, the older sibling was monitored through serial echocardiographic imaging. Throughout the 2.5-year follow-up period, there was a gradual deterioration in cardiac function, ultimately requiring HTx. The decline in cardiac function and dilatation of the LV is evident in echocardiographic images **(Figure 1B)**. The LVIDd increased from 48 mm to 58 mm (z-score 2.5) and LVEF measured 28%, assessed 5 months prior to HTx (**Figure 1B**, right panels). In line with the proband’s clinical presentation, the apparent deterioration in LV function was accompanied by a concomitant dilatation of the left atrium and a decline in right ventricular function. Throughout the follow-up period, NT-proBNP and hs-cTnT levels exhibited a drastic increase, which declined after an uncomplicated HTx procedure **(Supplemental Figure 1, C and D)**. Like the proband, the older sibling also exhibited moderate renal insufficiency, which slowly aggravated post-HTx, despite a tacrolimus-sparing immunosuppressive regimen. Additional clinical details are provided in **Supplemental Table 1**. Notably, analysis of the family history revealed that three cousins (individual III:4, III:5 and III:6; **Figure 1A**) died in early adolescence (age range: 11–15 years) from cardiac and renal failure. Unfortunately, medical records were not available.

### Histopathological examination of cardiac tissue from both siblings

Microscopic examination of the explanted hearts from both affected siblings revealed widespread fibrotic changes across the LV myocardium, consistent with the distribution of LGE on CMR **(Figure 2)**. Masson trichrome staining confirmed diffuse myocardial fibrosis in the proband **(Figure 2C)**. Furthermore, both siblings exhibited hypertrophy of cardiomyocytes in the LV myocardium cardiomyocyte disarray, especially in the proband. The septal myocardial changes were characterized by extensive fibrosis and hypertrophic cardiomyocytes in both individuals. Interestingly, examination of the right ventricle also revealed significant changes in myocardial tissue in this region. Cardiomyocyte hypertrophy was also visible in the right ventricle of both siblings, with interstitial fibrosis in the older sibling III:1 and more pronounced fibrosis in the proband.

**Figure 2.**
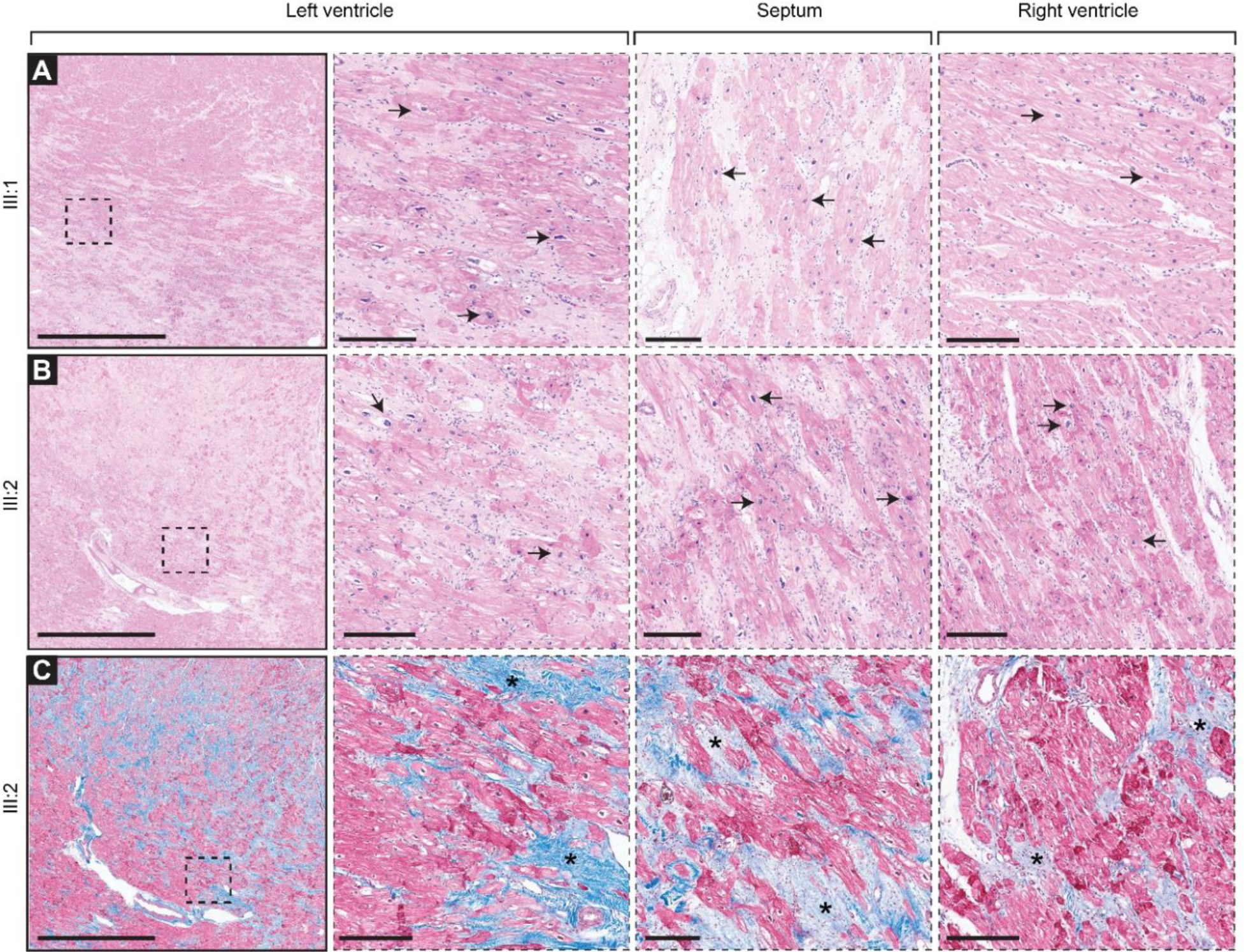
Histopathological assessment of explant-derived myocardial tissue. Top row: hematoxylin and eosin (HE) stain of myocardial tissue from the left ventricle (LV), septal wall and right ventricle (RV) of individual III:1. Middle row: HE stain of myocardial tissue from the LV, septal wall and RV of individual III:2. Bottom row: trichrome stain of LV, septal wall and RV of individual III:2. For the LV an overview and magnifications of the boxed regions are provided. Note the regions of fibrosis (asterisks) and hypertrophy of cardiomyocytes (arrows) in all regions. Scale bar overview, 2,5 mm; scale bar zooms ins: 250 μm.

### Genetic analysis detected a homozygous variant in the *XPNPEP3* gene

To determine the genetic cause for the observed phenotype, family-based exome sequencing (ES) revealed a homozygous variant c.1357G>A (chr22:41320486G>A, hg19), p.(Gly453Ser) in the *XPNPEP3* gene (NM_022098.4). This variant was also present in homozygous state in the older sibling III:1. The parents were heterozygous carriers, as was the healthy sibling III:3. No other likely pathogenic or pathogenic variants were identified **(Supplemental Table 2)**. To exclude the possibility that mtDNA variants contribute to the cardiac phenotype, we analyzed extracted mtDNA from cardiac tissue of the proband with next-generation sequencing. But, no known likely pathogenic or pathogenic variants (and/or large deletions) in the mtDNA genes were detected.

Expanding our investigation, linkage analysis was conducted using a fully penetrant recessive model, utilizing the single-nucleotide polymorphism markers derived from the exomes of both affected siblings and their parents (II:1, II:2, III:1, and III:2). Although low logarithm of the odds (LOD) scores were anticipated due to the limited numbers of family members included, the analysis revealed homozygous linkage peaks on chromosomes 2, 3, 4, 5 6, 8, 12, 16, 18, 19 and 22, with maximum LOD score of 1.78 **(Supplemental Figure 2)**. Notably, the identified variant in *XPNPEP3* located within one of the regions with the highest LOD scores, suggestive of genetic association.

To provide further genetic evidence linking *XPNPEP3* to cardiomyopathy, we aimed to identify additional families affected by the condition. Therefore, we reanalyzed ES data from two independent Dutch cardiomyopathy cohorts. Unfortunately, this analysis did not reveal any additional bi-allelic coding variants.

### The c.1357G>A variant in *XPNPEP3* results in aberrant splicing with loss-of-function (LoF) effect

Splice prediction algorithms anticipated that the c.1357G>A, p.(Gly453Ser) variant could impact RNA splicing by loss of a splice donor site and possible usage of an alternative splice donor site (c.1357+31). To validate this prediction, RNA was extracted from cultured skin fibroblasts from both affected siblings. Cells were cultured both with and without cycloheximide (CHX) to inhibit nonsense mediated mRNA decay (NMD). In control fibroblasts, only canonical PCR product was detected. Conversely, in both siblings only a larger product was detected **(Figure 3A and B)**. Subsequent Sanger sequencing analysis confirmed the use of the cryptic donor splice site in intron 9, resulting in 31-base pair (bp) intron retention inducing a frameshift p.(Gly453Serfs18*) **(Figure 3C)**. Notably, no normal transcript could be identified in both siblings. Treatment with CHX did not show differences compared to products from untreated cells, suggesting that mRNA degradation did not occur and mRNA is stable **(Figure 3, A and C**).

**Figure 3.**
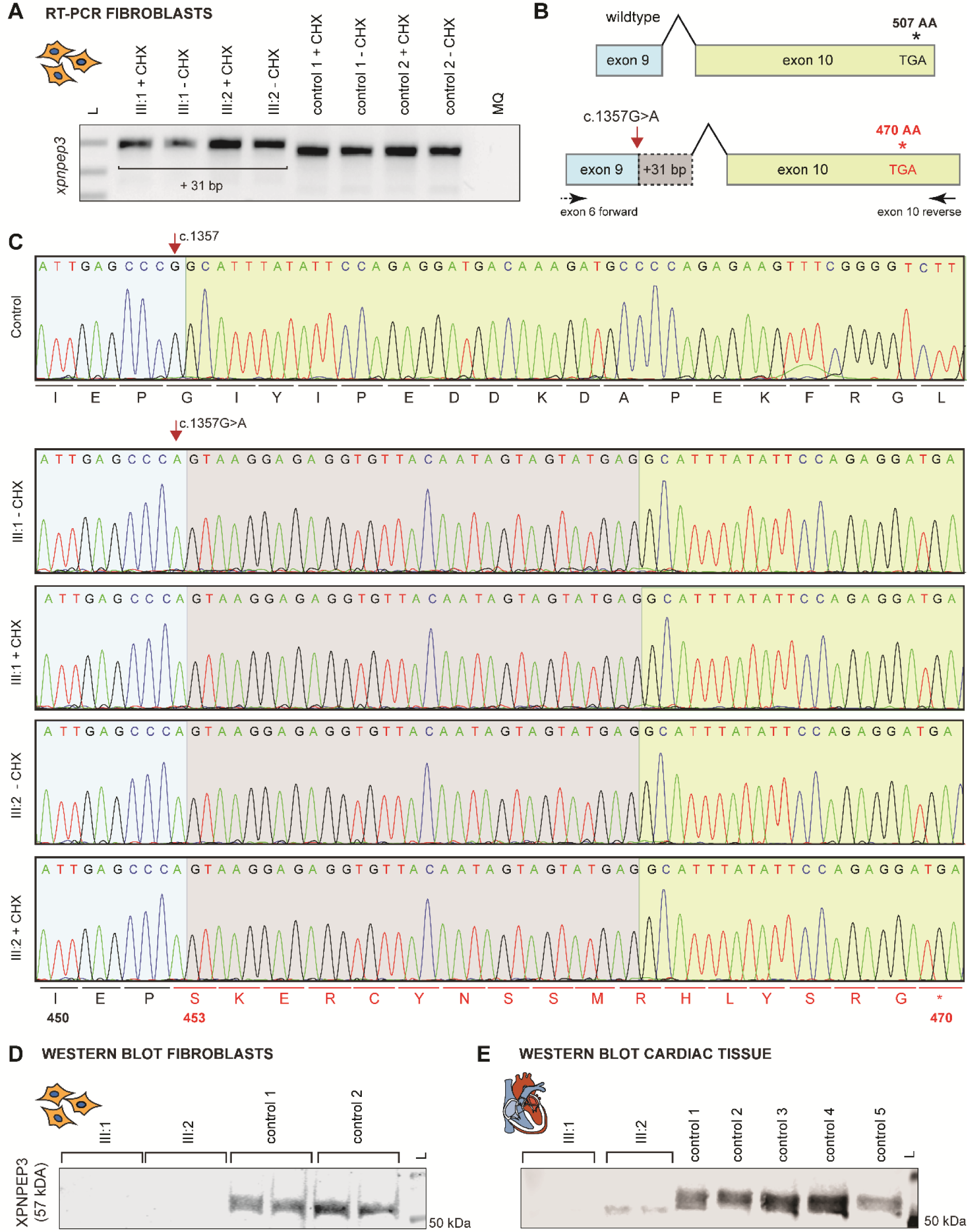
Impact of the *XPNPEP3* c.1357G>A variant on splicing and protein expression. (A) Gel electrophoresis of RT-PCR products of RNA isolated from biopsy-derived skin fibroblasts of both affected siblings and healthy controls that were cultured with and without cycloheximide (CHX). Note that the mutant product appears at a higher position on the gel due to the intron retention. L, ladder; MQ, Milli-Q water (B) Schematic overview of splice variant with use of an alternative splice donor site (c.1357+31). At the protein level this intron retention is anticipated to result in shorter protein due to an early stop codon at position 470. (C) Sequence analysis demonstrating use of an alternative splice donor site (c.1357+31) in cells from both siblings, leading to the 31-bp intron retention and frame shift. (D) Western Blot analysis of XPNPEP3 protein expression in fibroblasts of both affected siblings and age-matched controls. Fibroblasts were cultured in duplicate. (E) Western blot analysis of XPNPEP3 in left ventricular (LV) myocardial tissue samples. Sample of affected siblings were loaded in duplicate. Five age-matched control samples from independent cases were used for comparison. Note that both siblings show a near-complete absence of XPNPEP3 expression. Normalized quantification of XPNPEP3 and Stain-Free total protein images used as loading controls are shown in Supplemental in **Supplemental** Figure 3.

At the protein level, this 31-bp intron retention and resulting frameshift is predicted to produce a shorter protein due to a premature stop codon at amino acid 470. Intriguingly, Western blot analysis demonstrated absence of XPNPEP3 in fibroblasts of both affected siblings **(Figure 3D and Supplemental Figure 3)**. Expanding this analysis to cardiac tissue from both siblings, we again observed barely detectable levels of XPNPEP3 protein expression **(Figure 3E and Supplemental Figure 3)**. These results indicate that the splice variant causes a LoF effect, leading to loss of XPNPEP3 activity. Notably, in the siblings the band appeared at a slightly reduced molecular weight, which may correspond to the truncated form of XPNPEP3 (470 AA, 53.0 kDa). This implies that the mutant protein is unstable and prone to degradation or that there is reduced translation of the abnormally spliced transcript.

### *XPNPEP3* LoF variants result in interfamilial phenotypic variability

Splice and truncating variants in *XPNPEP3* have previously been associated with NPHPL1. However, recent reports suggest that the phenotypic spectrum is broader, including extra-renal manifestations [20–24]. In **Supplemental Table 3** and **Supplemental Figure 4** an overview of the reported variants and families is provided. Cardiomyopathy has been observed in three other children (age range: 6–15 years) from two affected families, referred to as family 3 and 6 in **Supplemental Table 3** and **Supplemental Figure 4** [20, 24]. All children also experienced renal failure. The cardiomyopathy in the proband of family 6 resulted in heart failure and death in early adolescence [24]. Interestingly, a nearly identical splice variant to that found in our family, has also been reported in three siblings from another family (age range: 16–30 years), referred to as family 2 in **Supplemental Table 3** and **Supplemental Figure 4**) experiencing renal problems, yet without a cardiac phenotype. This variant occurs at the same splice site as in our family, but involves a different nucleotide change (c.1357G>T instead of G>A). To test whether residual normal splicing could potentially explain phenotypic differences, a mini-gene exon trapping assay was performed in HEK 293T cells. An identical pattern of variant-induced mis-splicing was observed for both nucleotide variants with no residual canonical splicing **(Supplemental Figure 5)**. Together, these findings demonstrate that LoF variants in *XPNPEP3*, which are typically associated with renal problems, can also result in cardiomyopathy and/or neuromuscular symptoms, stressing the phenotypic heterogeneity associated with these variants.

### Loss of XPNPEP3 leads to structural and functional mitochondrial alterations in cardiac tissue

Assuming that XPNPEP3 is indeed the human ortholog of ICP55 – an essential mitochondrial matrix protein regulating mitochondrial function in yeast and plants – we next aimed to determine whether mitochondrial abnormalities could be detected. Therefore, mitochondrial respiratory chain enzyme activities were quantified in cardiac tissue from both siblings and compared to control tissues obtained from patients that had no underlying mitochondrial disease. This revealed an isolated deficiency in complex IV activity (cytochrome c oxidase), while the activities of complexes I-III remained within a control range **(Table 1)**. Of note, mitochondrial respiratory chain enzyme activities in cultured skin fibroblasts were within control ranges **(Supplemental Table 4)**.

**Table 1.** Mitochondrial respiratory chain enzyme activities in cardiac tissue from both siblings with novel homozygous splice variant in *XPNPEP3* c.1357G>A, p.(Gly453Ser)

|  | Activity<br>Individual III-1 | Activity<br>Individual III-2 | Control range | Unit |
| --- | --- | --- | --- | --- |
| Complex I | 219 | 312 | 77 – 646 | mU/UCII |
| Complex II | 196.5 | 258 | 65 – 150 | mU/mgprotein |
| Complex III | 1315 | 1017 | 1047 – 1841 | mU/UCII |
| Complex II+III (Succ.:cyt.c oxidoreductase; SCC) | 62 | 80 | 24 – 52 | mU/mg |
| Complex IV | 453 ↓ | 394 ↓ | 794 – 1396 | mU/UCII |

As an independent marker for mitochondrial disease, serum fibroblast growth factor (FGF21) in blood in both siblings was elevated: 6780 pg/ml in sibling III:1 (pre-HTx) and 264 pg/ml in III:2 (post-HTx); control range: 0-200.

Furthermore, transmission electron microscopy (TEM) was performed on LV myocardium to assess changes in mitochondrial morphology and cardiomyocyte architecture. To identify potential distinctive features linked to XPNPEP3 dysfunction, the acquired images were compared to previously generated TEM images of myocardial samples from a pediatric non-failing (NF) donor, an explanted heart of a pediatric patient with idiopathic DCM, and a myectomy sample from a pediatric patient with obstructive HCM **(Figure 4)**. A range of ultrastructural alterations in the myocardium of both siblings was observed compared with the NF donor. Disrupted cardiomyocyte architecture was apparent from a loss of myofibrillar area and abnormal clustering of mitochondria. Additionally, individual mitochondria in the tissue of both siblings typically displayed disrupted cristae structure, appearing fragmented and tubular rather than lamellar. Mitochondria often appeared large and swollen with a reduced density of cristae. Notably, mitochondria in cardiac tissue of both siblings seemed to be shaped almost exclusively spherical rather than elongated. Lastly, a unique and striking feature of the myocardium upon loss of XPNPEP3 was the ubiquitous presence of vacuole-like structures filled with electron-dense multi-layered hyaline bodies.

**Figure 4.**
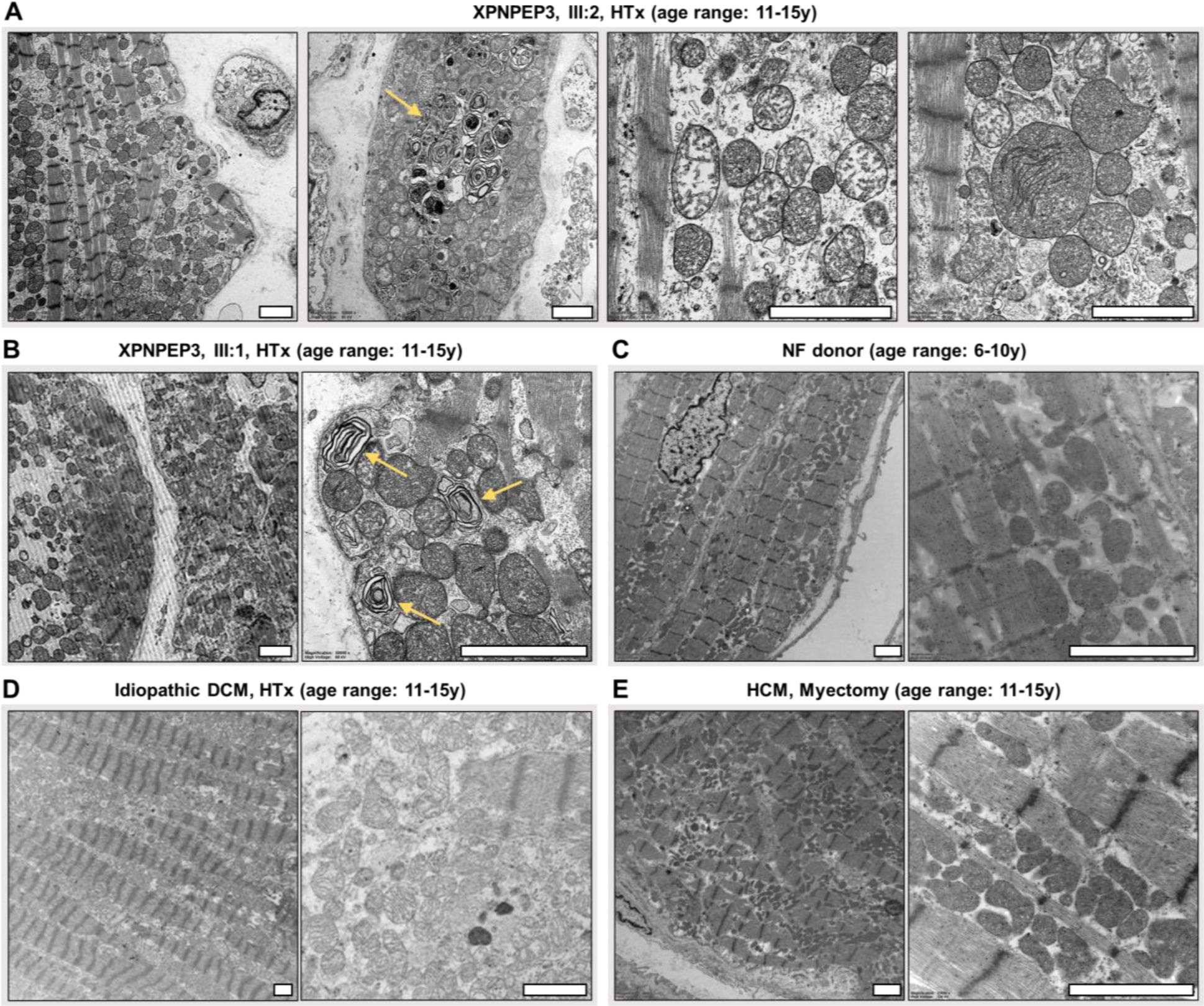
Transmission electron microscopic (TEM) images of patient-derived cardiac tissue from both siblings compared to various controls. (A) LV myocardial tissue derived from sibling III:2 upon heart transplantation (HTx) (age range: 11–15 years). (B) LV myocardial tissue derived from sibling III:1 upon HTx (age range: 11–15 years). (C) LV myocardial tissue from a non-failing (NF) donor (age range: 6–10 years). (D) LV myocardial tissue from an individual with idiopathic dilated cardiomyopathy (DCM) upon HTx (age range: 11–15 years). (E) Myectomy sample of the LV myocardium from an individual with obstructive hypertrophic cardiomyopathy (HCM) (age range: 11–15 years).

### Modelling Xpnpep3 deficiency in zebrafish

To gain insight into the role of XPNPEP3 in mitochondrial function and disease, we used zebrafish as vertebrate model system, renowned for its effectiveness in modeling human diseases and investigating cardiac and mitochondrial functions [37–40]. The zebrafish Xpnpep3 homolog encodes a 510-amino acid (AA) protein with 57% sequence similarity to the human XPNPEP3 protein (507 AA). Computational 3D modelling confirmed structural conservation, suggesting functional preservation **(Supplemental Figure 6)**. To evaluate the loss of XPNPEP3 function in vivo and model the LoF mechanism, we used CRISPR/Cas9 genome editing to endogenously target the zebrafish *xpnpep3* locus **(Figure 5A)**. A 7 bp deletion was introduced in exon 9 of the gene, resulting in a predicted shorter protein due to an early stop codon. Consistent with observations in the patients, in homozygous mutant larvae (referred to as *xpnpep3^Δ7/Δ7^*) we detected truncated *xpnpep3* mRNA with the 7 bp deletion **(Figure 5B)**. Unfortunately, we did not find a good working antibody for zebrafish, leaving us uncertain about the presence or absence of the mutated protein (results not shown). Homozygous mutant larvae were obtained at the expected Mendelian ratio, and showed no morphological differences from wild-type (*xpnpep3^+/+^*) and heterozygous (*xpnpep3^+/Δ7^*) siblings up to 5 days post fertilization (dpf) **(Figure 5C and D)**. More specifically, the pronephros exhibited normal morphology without renal cysts. Also, cardiac morphology, ventricular dimensions and cardiac performance were within normal limits, with no evidence of cardiac oedema **(Figure 5C-F)**. During the initial visual examination of the zebrafish larvae prior to the experiments described above, *xpnpep3^Δ7/Δ7^*mutants appeared less active and exhibited a lethargic behaviour compared to their wild-type siblings. However, this was not a consistent feature. Noteworthy, touch-response behavior was normal, indicating no loss of sensory-motor function **(Figure 5G)**.

**Figure 5.**
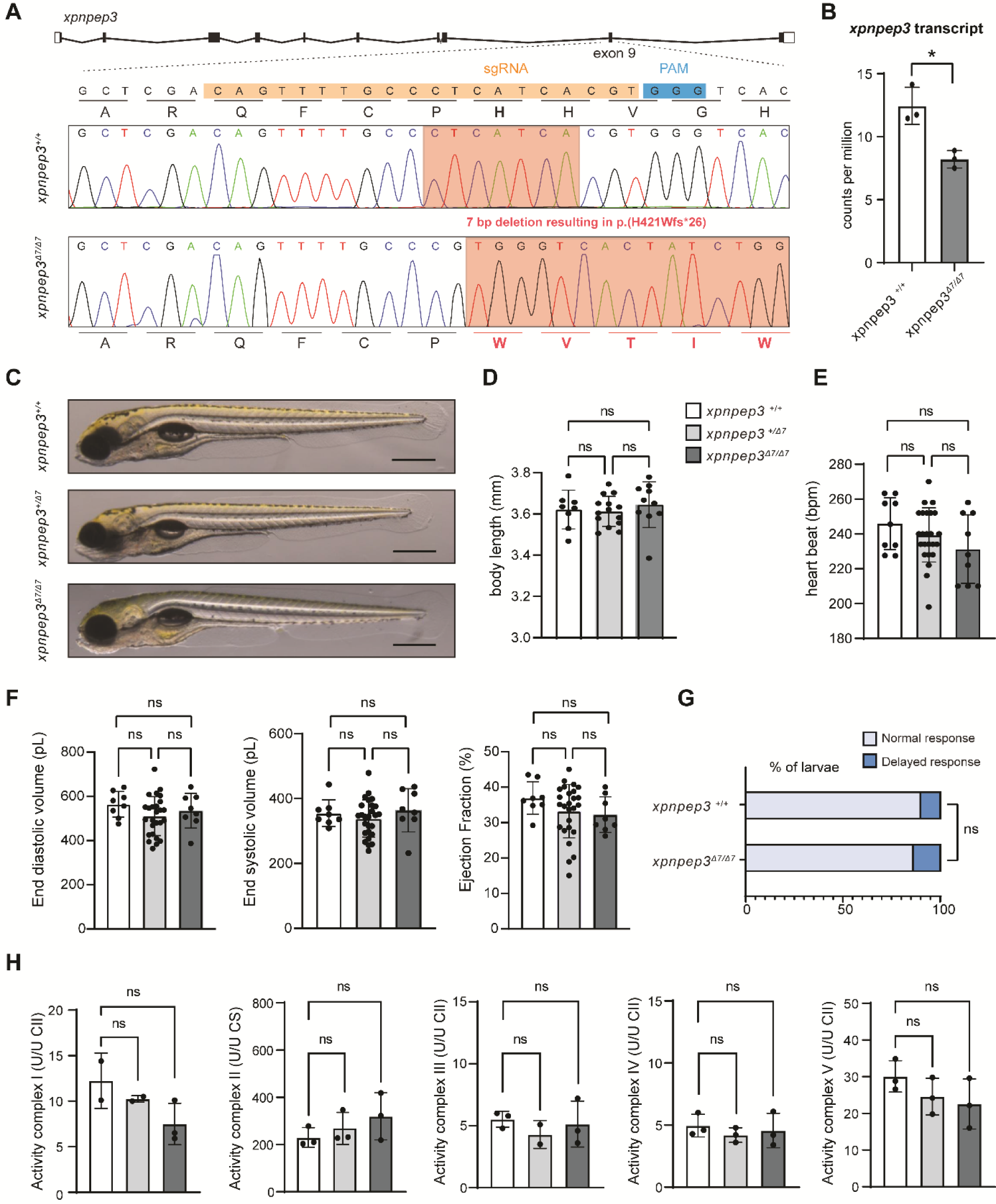
*xpnpep3^Δ7/Δ7^*zebrafish have normal cardiac function and mitochondrial activity during early development. (A) CRISPR/Cas9 strategy used for targeted disruption of *xpnpep3* inducing a 7 bp deletion in exon 9, resulting in a frameshift. On protein level this frameshift is anticipated to lead to a truncated protein shorter than the wild-type due to a premature stop. (B) Graph showing *xpnpep3* mRNA total read counts (counts per million, CPM) from RNA-sequencing data **(see** Figure 6) of 5 days post-fertilization (dpf) zebrafish larvae. *n*=3 biological replicates per genotype. Unpaired t-test was used to test for significance. *p<0.05 was considered significant. (C) Representative images of 4 dpf larvae, illustrating no gross morphological abnormalities compared to heterozygous and wild-type siblings. Note that there are no signs of cardiac oedema nor renal cysts. (D) Body length measurements of larvae at 4 dpf, with *n*=8 for *xpnpep3^+/+^, n=14 for xpnpep3^Δ7/+^ and n=10 for xpnpep3^Δ7/Δ7^* Data are represented as mean ± SD, and one-way ANOVA coupled with Tukey’s multiple-comparison test was used to test for significance. (E-F) Ventricular contractility parameters derived from high-speed imaging movies of 5 dpf larvae, including heartbeat, end-diastolic volume, end-systolic volume, and ejection fraction, with *n=*8 for *xpnpep3^+/+^, n=*26 for *xpnpep3^Δ7/+^* and *n*=8 for *xpnpep3^Δ7/Δ7^*. Data are displayed as mean ± SD; one-way ANOVA coupled with Tukey’s multiple-comparison test was used to test for significance. (G) Touch response assay of 3 dpf larvae, with *n=20* for *xpnpep3^+/+^* and *n*=10 for *xpnpep3^Δ7/Δ7^* Fisher’s exact test was used to test for significance. (H) Enzymatic activities of the respiratory chain complexes were assessed in lysates from pooled larvae of 5 dpf (*n=*80 per genotype). Measurements were performed in triplicate, but due to some failures, only duplicate measurements were available for certain samples. Data were normalized to the activity of Complex II (expressed as units per unit of complex II activity, U/U CII) and are presented as mean ± SD. Complex II was normalized to the activity of Citrate synthase (CS). Kruskal-Wallis test with Dunn’s multiple comparisons test was used to test for significance. p<0.05 was considered significant.

Given the mitochondrial findings in the affected siblings, we next determined whether similar indications could be identified in the zebrafish. The activities of mitochondrial respiratory chain enzymes were measured using pooled samples of 5 dpf whole-body larvae, but no statistically significant differences were detected **(Figure 5H)**.

To detect a possible synthetically induced phenotype through stress, we also tested several stress conditions. Larvae were grown at elevated temperatures (32°C compared to 28°C) to assess the impact of thermal stress. We also added the beta-adrenergic agonist isoproterenol to the water to induce cardiac stress by increasing heart rate. Additionally, we administered mitochondrial complex inhibitors to the water, including rotenone (complex I inhibitor), sodium azide (complex IV inhibitor), and oligomycin (complex V inhibitor), to disrupt mitochondrial function. We tested different concentrations at various time points and durations to assess their effects. Despite these interventions, no conditions were identified that selectively induced the phenotype in the mutants while leaving the wild-type unaffected (results not shown).

### Transcriptomic and proteomic analysis of *xpnpep3^Δ7/Δ7^* zebrafish reveals coordinated expression differences impacting mitochondrial function

Despite the absence of an observable disease phenotype in early zebrafish development, our next objective was to investigate potential early dysregulation of molecular processes and pathways resulting from the loss of wild-type Xpnpep3 function. To do so, we isolated total RNA and proteins from *xpnpep3^Δ7/Δ7^*mutants and wild-type siblings at 5 dpf **(Figure 6A)**. RNA sequencing (RNA-seq) and quantitative mass spectrometry-based proteomics analyses were used to identify differently expressed genes (DEGs) and differentially expressed proteins (DEPs) between *xpnpep3^Δ7/Δ7^* larvae and their corresponding wild-type siblings. DEGs were based on a false discovery rate (FDR)-corrected p-value of <0.05, and an absolute log2 fold-change (log2FC) of > 1.0-fold. DEPs were determined using a p-value of <0.05 and an absolute log2FC> 1.0-fold. The wild-type and *xpnpep3^Δ7/Δ7^* RNA-seq and mass spectrometry data clustered independently suggesting the presence of changes related to the genotype **(Supplemental Figure 7)**. Differential analysis showed changes of 189 DEGs and 541 proteins DEPs, of which 115 DEGs and 400 DEPs were downregulated, and 74 DEGs and 151 DEPs were upregulated **(Figure 6B-D, and Online Supplemental Data files 1 and 2).** Of note, the Xpnpep3 protein could not be detected in proteomic analysis for both the mutant or wild-type siblings, most likely due to its relatively low absolute abundance in the proteome.

**Figure 6.**
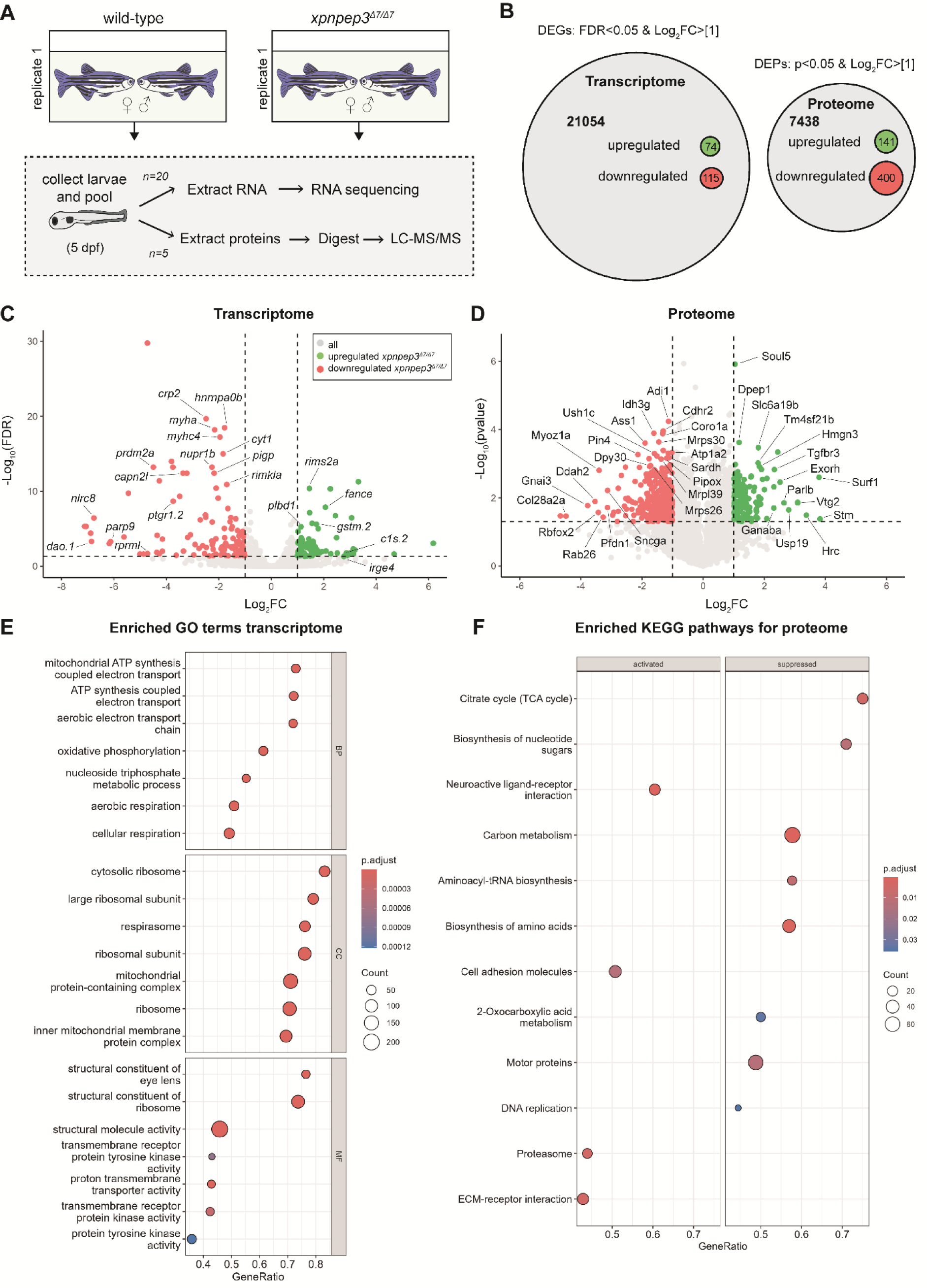
Global transcriptomic and proteomic analysis in *xpnpep3^Δ7/Δ7^* zebrafish reveals mitochondrial changes. Transcriptomic and proteomic analysis comparing genes and proteins quantified in *xpnpep3^Δ7/Δ7^*zebrafish mutants relative to wild-types at 5 days post fertilization (dpf). (A) Schematic presentation of the experimental design. At 5 dpf, *xpnpep3^Δ7/Δ7^*and wild-type larvae were collected. Per genotype, for each of the three biological replicates (offspring from 3 different breedings), *n=*20 larvae were pooled together for RNA extraction and *n=*5 were pooled together for protein extraction. (B) Proportional area chart representing total numbers of genes and proteins identified through RNA sequencing (RNA-seq) and quantitative mass-spectrometry analysis. The number of differentially expressed genes (DEGs) was determined based on a false discovery rate (FDR)-corrected p-value <0.05, and an absolute log2FC > 1.0-fold. The number of differentially expressed proteins (DEPs) was determined using a p-value of <0.05 and an absolute log2FC > 1.0-fold. Upregulated DEGs and DEPs are idicated in green and the dowregulated ones are indicated in red. (C) Volcano plot representation of transcriptomic data. Green and red dots denote the DEGs, the gray dots represent genes that were not differentially expressed. The log2FC is represented on the x-axis, and the -log10 FDR of these differences is plotted on the y-axis. DEGs with the most notable significance and log2FC are labelled. DEGs lacking clear gene annotation were omitted from labeling. (D) Volcano plot representation of proteomic data. Green and red dots denote the DEPs, gray dots represent proteins that were not differentially expressed. The x-axis represents log2FC in protein quantity, while the y-axis shows the -log10 p-value of these changes. Proteins with the most notable significance and log2FC are labelled. DEPs lacking clear annotation were omitted from labeling. (E) Dot plot visualization of the Gene Set Enrichment Analysis (GSEA) on the RNA-seq data using the Gene Ontology (GO) classification. The x-axis shows ratio of genes to number of genes in the term. The size of each circle denotes the counts of genes in the term and colour denotes adjusted p-value. The top term biological processes (BP), cellular compartment (CC) and molecular function (MF) are shown. (F) Dot plot visualization of GSEA-derived enriched Kyoto Encyclopedia of Genes and Genomes (KEGG) pathways in proteomic data.

We conducted Gene Set Enrichment Analysis (GSEA) using the Gene Ontology (GO) database to determine significantly overrepresented gene classes **(Figure 6E, Supplemental Figure 8A, and Online Supplemental Data file 3)**. Most significantly affected biological processes were all related to mitochondrial energy production and metabolic functions (e.g. mitochondrial ATP synthesis and coupled electron transport, oxidative phosphorylation, and cellular respiration). Key cellular components included ribosomal and mitochondrial structures, involved in mitochondrial protein synthesis, respiration and function. Top molecular function included activities needed for signal transduction, membrane transport, and maintaining structural integrity **(Figure 6E)**. These findings indicate that the genes exhibiting the most pronounced coordinated expression differences between *xpnpep3^Δ7/Δ7^* and wild-type conditions predominantly regulate mitochondrial energy metabolism, and are also involved in the production of mitochondrial proteins, including their synthesis, folding and proper localization.

The Kyoto Encyclopedia of Genes and Genomes (KEGG) and Reactome databases were used to perform pathway enrichment analysis on the proteomic dataset. This analysis showed that the citric acid (TCA) cycle, the central mitochondrial metabolic pathway, was most significantly suppressed in *xpnpep3^Δ7/Δ7^*zebrafish using both databases **(Figure 6F and Supplemental Figure 9A)**. Additionally, biosynthesis pathways such as carbon, nucleotide sugars, amino acids, and aminoacyl tRNA were also enriched and suppressed. Conversely, top enriched activated pathways included amongst others neuroactive-ligand mediated neurotransmitter interactions, cell-adhesion, proteasome, autodegradation of the E3 ubiquitin ligase COP1, and stabilization of p53. The enrichment of these diverse pathways not only implicates mitochondrial dysfunction, but also the involvement of other cellular metabolic changes and an increased cellular response to stress.

We also investigated the proteomic dataset using the GO database and visualized it using a Treemap **(Supplemental Figure 8B and 9B)**. In addition, the transcriptome data was also analysed for enrichment of KEGG and Reactome pathways **(Supplemental Figure 9, C and D)**. These analyses had similar outcomes as recurrent findings included mitochondrial suppression, accompanied by alterations in metabolic pathways, DNA replication, ribosome biogenesis, RNA processing, and protein synthesis, including folding and degradation. Interestingly, transcriptome analysis identified cardiac muscle contraction (KEGG: dre04260) as an enriched suppressed pathway and the calcium signalling pathway (KEGG: dre04020) as an activated one **(Supplemental Figure 9D)**.

### Decreased mitochondrial protein abundance in *xpnpep3^Δ7/Δ7^* zebrafish

Based on XPNPEP3’s suggested role in mitochondrial protein processing in yeast and plants and our consistent omics analysis results indicating its primary impact on mitochondrial function in the zebrafish, we specifically focused on mitochondrial associated proteins. To do so, we utilized a curated list known as the “MitoCarta Inventory”, which catalogs genes encoding mitochondrial proteins playing diverse role within the organelle. In total, the list contained 1326 entries, of which 1056 could be mapped to the transcriptomic dataset and 680 to the proteomics dataset **(Figure 7A, Online Supplemental Data file 4)**. On the individual transcript level, there is no robust effect: among all genes examined, only one gene, alanyl-tRNA synthetase 2 (*aars2*), was identified as a DEG (FDR <0.05, absolute LogFC >1) **(Figure 7, A and B)**.

**Figure 7.**
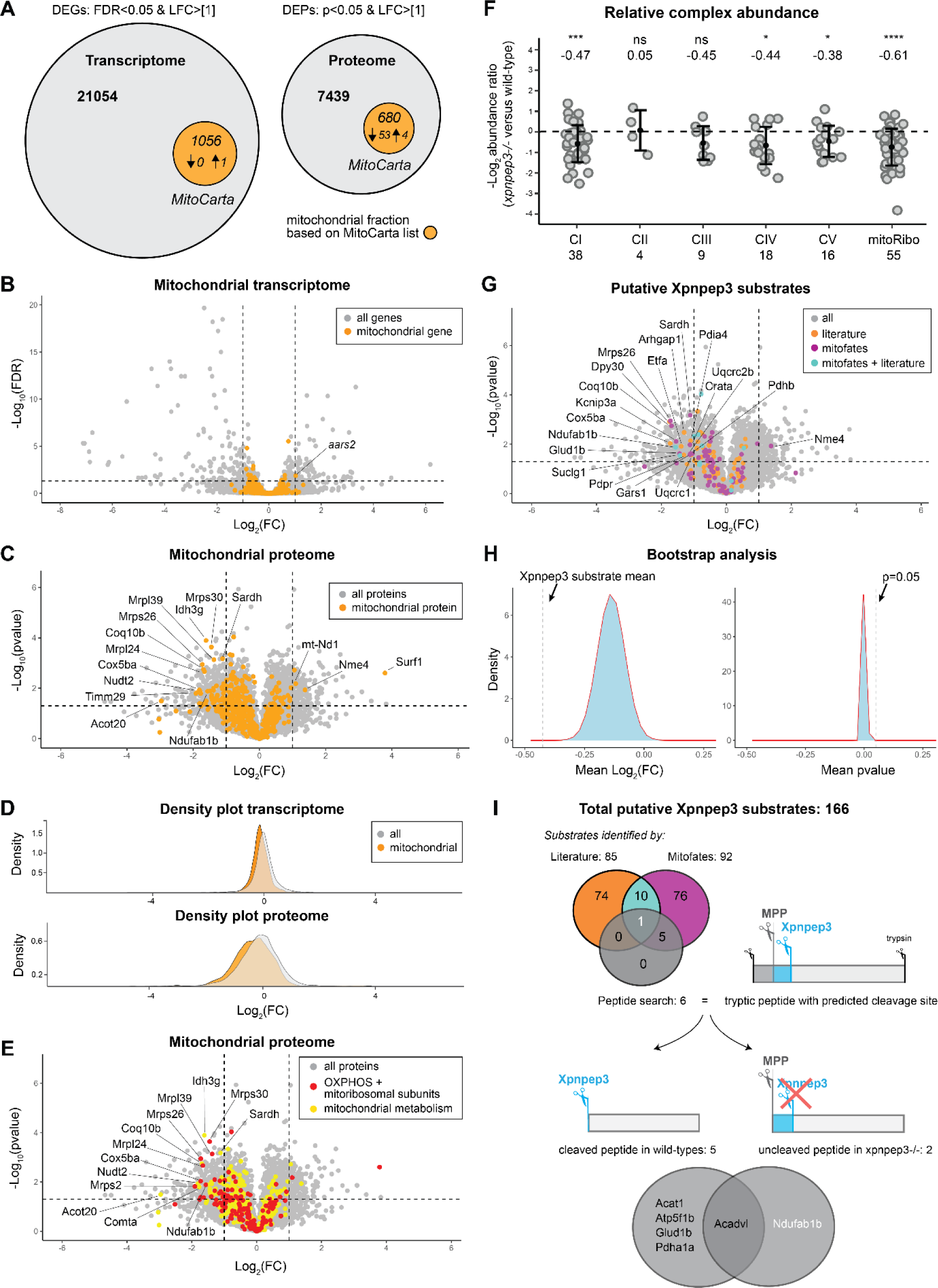
Mitochondrial proteins (including putative Xpnpep3 substrates) show lower abundance in *xpnpep3^Δ7/Δ7^* zebrafish. Transcriptomic and proteomic analyses comparing mitochondrial genes and proteins quantified in *xpnpep3^Δ7/Δ7^* zebrafish mutants relative to wild-types at 5 dpf. (A) Proportional area chart illustrating the proportionate number of genes and proteins identified through RNA-seq and quantitative mass-spectrometry analysis. The orange circle represents the mitochondrial fraction that was filtered in silico using the MitoCarta annotation (the *Danio rerio* MitoCarta list used is provided in **Online Supplemental Data file 4**). Within the mitochondrial fraction, the number of DEGs and DEPs are indicated. DEGs were based on a FDR-corrected p-value<0.05, and an absolute log2FC > 1.0-fold. DEPs were determined by a p-value of <0.05 and an absolute LogFC > 1.0-fold. The direction of the arrow in the graphical representation indicates whether the DEGs/DEPs are up- or downregulated. (B) Volcano plot representing the complete transcriptome, with mitochondrial genes highlighted in orange atop of other genes. (C) Volcano plot depicting the complete proteome, with mitochondrial proteins highlighted in orange atop of other proteins. Well-annotated DEPs with most notable significance and log2FC were labelled. (D) Density plot visualizing distribution of log2FC of mitochondrial genes/proteins in comparison to the density of the complete transcriptome/proteome. (E) Volcano plot depicting the proteome with a subset of mitochondrial proteins highlighted categorized according to the MitoCarta annotation. (F) The relative complex abundance (RCA) plot of the OXPHOS complexes and mitoribosome. The graph represents each complex ratio of *xpnpep3^Δ7/Δ7^*zebrafish mutant versus wild-types with ± standard deviation. The mean ratio of each protein within a complex is calculated, and a ratio paired t-test was performed to determine significance of each complex. p<0.05 was considered significant. (G) Volcano plot of the complete proteome with putative Xpnpep3 substrates based on literature review coloured in orange, putative substrates based on the mitofates prediction tool [43] in purple, and substrates that were based on both shown in light blue. Of these substrates, the ones showing differential expression, determined by using a p-value of <0.05 and an absolute log2FC > 1.0-fold, were labelled. (H) The left graph shows the distribution curve of the mean log2FC of 100.000 randomly selected protein sets (containing n=146 proteins, which contains the same number of proteins as the Xpnpep3 substrate group included in statistical analysis). The right graph depicts the p-value distribution of a one-tailed Student-t test comparing the log2FC between the Xpnpep3 substrate group and each respective random protein set. (I) Peptide-based analysis revealing six high-confidence putative substrates of Xpnpep3.

In the mitochondrial protein analysis, 57 mitochondrial proteins were identified as DEPs, with 53 showing downregulation and 4 showing upregulation **(Figure 7, A and C)**. The mitochondrial DEPs, involved in various mitochondrial processes and localized within distinct mitochondrial subcompartments, exhibit lower abundance compared to all identified proteins, as indicated by a leftward shift in abundance levels observed in both volcano and density plots **(Figure 7C-E, Supplemental Figure 10, Online Data Supplement File 5)**. The majority of the downregulated DEPs participate in mitochondrial metabolism **(Figure 7E)**. Next to this, mitochondrial DEPs also encode OXPHOS subunits, important for electron transport chain and ATP synthase function, and mitochondrial ribosomal subunits, involved in mtDNA-encoded protein synthesis. Interestingly, surfeit locus protein 1 (Surf1), which is reported to be involved in correct assembly and function of complex IV, is the most upregulated DEP [41]. To further understand the composition of OXPHOS complexes and mitochondrial ribosomes, we also plotted the relative complex abundance (RCA) of all OXPHOS and mitoribosomal proteins as a ratio between *xpnpep3^Δ7/Δ7^* and wild-types. This revealed a significantly reduced abundance of OXPHOS complexes I, IV, V, and the mitochondrial ribosomes in *xpnpep3^Δ7/Δ7^* zebrafish **(Figure 7F)**. Taken together, these findings show that the integrity of the mitochondrial proteome is affected and many important mitochondrial proteins involved in mitochondrial metabolism, respiration, and protein synthesis, are lower expressed in the *xpnpep3^Δ7/Δ7^* larvae during early development.

### Putative Xpnpep3 substrates are significantly less abundant in *xpnpep3^Δ7/Δ7^* zebrafish

We next aimed to identify potential Xpnpep3 substrates whose improper processing may contribute to this proteomic reduction. To accomplish this, we first compiled a list of putative Xpnpep3 targets which was based on experimentally determined ICP55 substrates in Saccharomyces cerevisiae and Arabidopsis thaliana [26, 27, 42] and putative XPNPEP3 substrates in humans [20]. For each of these proteins, we manually selected its Danio rerio ortholog. Furthermore, we utilized the Mitofates prediction tool to analyze the entire Danio rerio proteome, enabling us to predict the presence of a mitochondrial presequence, a MPP and ICP55/XPNPEP3 cleavage site for every protein tested [43]. In total, we identified 166 proteins as putative substrates of Xpnpep3: 85 from literature research and 92 from computational predictions using Mitofates, with 11 substrates predicted by both **(Online Supplemental Data file 5)**. Out of these substrates, 146 had measurable quantities in at least 5 out of 6 replicates and were included in our statistical analysis **(Figure 7G)**. Interestingly, some of these proteins, including sarcosine dehydrogenase (Sardh), mitochondrial ribosomal protein S26 (Mrps26), cytochrome c oxidase subunit 5b (Cox5ba; complex IV) and NADH:ubiquinone oxidoreductase subunit AB1b (Ndufab1b; complex I), are among the most significantly downregulated mitochondrial DEPs in *xpnpep3^Δ7/Δ7^* zebrafish **(Figure 7C)**.

To test whether this group of substrates was significantly lower in abundance in *xpnpep3^Δ7/Δ7^* zebrafish, we conducted a bootstrap analysis by randomly drawing 100,000 protein sets of the same size (n=146) from our proteomics dataset. For each iteration, the mean log2FC of these sets was calculated and a one-tailed Student’s t-test was performed **(Figure 7H)**. The putative substrate group exhibited a mean log2FC of -0.424 ± 0.056, which was lower than the mean of the bootstrap distribution (-0.138 ± 0.056). The resulting mean p-value was 0.002 ± 0.007, indicating strong evidence of downregulation. Together these results demonstrate that the putative Xpnpep3 substrates in the *xpnpep3^Δ7/Δ7^*mutants are significantly less abundant compared to any randomly selected same-sized protein set. This strongly suggests that the reduced abundance of these substrates is the result of Xpnpep3 deficiency.

Of the 166 putative substrates, 20 were unsuitable for statistical analysis and were consequently not represented in the volcano plot **(Supplemental Figure 11)**. Yet, the ones showing stable expression in wild-type, but minimal to no expression in *xpnpep3^Δ7/Δ7^* replicates, are still of significant interest, as this can be explained by Xpnpep3 dysfunction. Particularly intriguing are the proteins that show expression in all three wild-type replicates but are absent in the *xpnpep3^Δ7/Δ7^*replicates, which include: thiosulfate sulfurtransferase like domain containing 1 (Tstd1), carbamoyl phosphate synthetase (Cps1), and GTP dependent ribosome recycling factor mitochondrial 2 (Gfm2), which are involved in mitochondrial metabolism and mitochondrial protein translation.

### Peptide-based analysis reveals high-confidence putative substrates of Xpnpep3

To investigate whether these substrates were improperly processed in *xpnpep3^Δ7/Δ7^* zebrafish, we conducted additional searches focused on N-terminal processed peptides within our proteomic dataset. Specifically, our analysis searched for protein N-terminal peptides with a conventional trypisin cleavage site (K/R) at the C-terminus, allowing up to 3 missed cleavages and derived from the first 100 amino acids of protein sequences. Despite limited detection of N-terminal peptides in our dataset, we next refined our search to include only the peptides whose N-termini matched the predicted Xpnpep3 cleavage site or were positioned one amino acid upstream. In wild-type zebrafish, we found peptide-based evidence of functional Xpnpep3 for 5 proteins, meaning that for these proteins only the processed peptide could be detected: mitochondrial lipid metabolism proteins acyl-CoA dehydrogenase very long chain (Acadvl), acetyl-CoA acetyltransferase 1 (Acat1), TCA cycle proteins glutamate dehydrogenase 1b (Glud1b) and pyruvate dehydrogenase E1 subunit alpha 1a (Pdha1a), and OXPHOS complex V subunit ATP synthase F1 subunit beta (Atp5f1b) **(Figure 7I and Online Supplemental Data file 6)**. Additionally, solely the unprocessed peptide of Acadvl could be detected in *xpnpep3^Δ7/Δ7^*zebrafish. Similarly, for Ndufab1b we could only detect the unprocessed peptide in *xpnpep3^Δ7/Δ7^* zebrafish. Interestingly, in the differential expression analysis Acadvl (p=0.005, log2FC =-0.669), Acat1 (p=0.058, log2FC =-1.533), Pdha1a (p=0.021, log2FC =-0.634), Atp5f1b (p=0.091, log2FC =-1.055), Ndufab1b (p=0.013, log2FC =-1.566) all exhibit downregulation as evidenced by their negative log2FC values. Only Ndufab1b, predicted to be an Xpnpep3 substrate by both Mitofates and experiments in yeast, was regarded as DEP using the stringent threshold criteria of p<0.05 and log2FC <-1 **(Figure 7C and G)**. All other proteins satisfy the criteria of p<0.1 and log2FC <−0.6, indicating that while their downregulation is notable it does not meet the criteria for significance.

Taken together, our peptide analysis provides evidence supporting mitochondrial proteins Acadvl, Acat1, Glud1b, Pdha1a, Atp5f1b, and Ndufab1b as putative specific substrates of zebrafish Xpnpep3. We have particularly high confidence in Acadvl and Ndufab1b because, under mutant conditions, only the unprocessed peptide was detectable. Additionally, Ndufab1b emerged as one of the DEPs in *xpnpep3^Δ7/Δ7^* zebrafish.

In summary, these findings demonstrate that Xpnpep3 functions as a regulator of the mitochondrial proteome in zebrafish. Its deficiency leads to subclinical mitochondrial pathology in zebrafish larvae, characterized by a significant reduction in the expression of various mitochondrial proteins. This reduction is likely due to the failure of Xpnpep3 to cleave destabilizing N-terminal residues from their peptide sequence. These findings provide further evidence that XPNPEP3 functions as the vertebrate ortholog of ICP55, playing a role in processing mitochondrial precursor proteins and maintaining mitochondrial health.

## Discussion

In this study, we provide clinical, genetic, and zebrafish disease modeling evidence revealing *XPNPEP3* as a causative gene for pediatric cardiomyopathy through a mechanism involving modulation of the mitochondrial proteome. We present the third family described in the literature known to be affected by *XPNPEP3*-related cardiomyopathy. Molecular analysis in this family identified a novel splice variant which resulted in loss of XPNPEP3 expression. Importantly, both affected children underwent cardiac transplantation for end-stage cardiac failure, illustrating the critical function of XPNPEP3 in cardiac health. Cardiac tissue analyses revealed mitochondrial dysfunction in both affected siblings, defined by ultrastructural abnormalities and reduced complex IV activity. Complementarily, we established a zebrafish model demonstrating subclinical mitochondrial pathology, characterized by suppression of specific gene sets associated with mitochondrial function. Proteomic profiling in these mutants revealed a notable reduction in the expression of mitochondrial proteins, including putative Xpnpep3 substrates, that carry out various functions within the mitochondria. Together, these findings strongly indicate that XPNPEP3 functions as the vertebrate ortholog of ICP55, involved in the processing and stabilization of mitochondrial precursor proteins and maintaining mitochondrial health. In summary, our study highlights the need to include *XPNPEP3* in diagnostic gene panels for mitochondrial and cardiac diseases. Given that *XPNPEP3*-related phenotypes can be associated with life-threatening cardiac manifestations, we propose regular cardiac evaluation in patients harboring bi-allelic LoF *XPNPEP3* variants. Furthermore, we emphasize the need to expand the patient cohort to comprehensively understand the disease etiology associated with this disease.

To evaluate the cause of early-onset heart failure in this family, CMR was performed in individual III:1, which showed hypertrophy and mid-wall and epicardial LGE alongside elevated native T1 and T2 times and ECV. Histopathological examination of cardiac tissue revealed diffuse fibrosis. Although non-specific, these findings may indicate mitochondrial dysfunction, where the accumulation of oxidative damage drives cardiac structural remodeling, such as hypertrophy and myocardial fibrosis [44–46]. To further explore the role of mitochondria in disease, we next compared myocardial TEM images from both siblings with those from pediatric cardiac tissue samples from a non-failing donor, as well as cases of DCM and obstructive HCM, which are rare and valuable samples, making this analysis of particular importance. The TEM images of both siblings revealed enlarged, spherical mitochondria with fragmented tubular cristae, which is a hallmark of mitochondrial cardiomyopathy [47–52]. The multi-layered hyaline bodies have also been reported in other mitochondrial cardiomyopathies [47], but are rarely observed in conditions like HCM, where mitochondrial dysfunction is secondary to sarcomere mutations, disrupted cardiomyocyte architecture and metabolic stress [53]. These results reinforce the hypothesis that the cardiac failure observed in patients with bi-allelic LoF *XPNPEP3* variants is caused by a primary mitochondrial defect. Additionally, functional testing of the respiratory chain enzyme activities in cardiac tissue of both siblings showed reduced complex IV activity. In contrast, O’Toole et al. reported an isolated complex I deficiency in skeletal muscle and cultured fibroblasts from one of the families exhibiting end-stage kidney disease [20]. One plausible explanation for this discrepancy could be that different tissues have distinct sensitivities and compensation mechanisms for mitochondrial problems [54–56]. Although invasive, this question could be addressed by testing respiratory chain enzyme activities in skeletal muscle biopsies from additional patients. Interestingly, other studies have shown faster turnover rates of COXI subunits of complex IV in mouse heart mitochondria compared to other subunits [57]. Consequently, complex IV could be more vulnerable for defective synthesis or assembly of OXPHOS subunits in relation to XPNPEP3 dysfunction in the heart, which may be an explanation for the isolated complex IV deficiency observed in our siblings.

To further understand the function of XPNPEP3 and to link its dysfunction with the disease, we generated a zebrafish model with a LoF mutation in *xpnpep3*. While we did not detect clear disease phenotypes in early zebrafish development, we observed mitochondrial alterations on transcriptomic and proteomic levels. Specifically, GSEA identified mitochondrial pathways with differential expression between wild-type and mutant conditions that were overlooked when only considering individual genes with the most significant expression differences. In particular, GSEA indicated that processes related to mitochondrial respiratory function, mitochondrial metabolism and mitochondrial translation machinery were suppressed in *xpnpep3^Δ7/Δ7^* larvae. Building on these results, we observed that the majority of mitochondrial proteins involved in these various mitochondrial functions showed downregulation, with a subset surpassing our threshold set for differential expression. Overall, we believe that the lack of pronounced changes in log2FC correlate with the absence of severe disease phenotypes in these larvae. We propose that these subtle changes are contributing factors to the initial pathological events as a result of Xpnpep3 dysfunction. Hence, it will be of interest to determine if disease phenotypes related to mitochondrial disease, in particular cardiomyopathy, will manifest in these mutants at later stages (>5 dpf). It will also be interesting to examine whether the mitochondrial molecular changes and stress response pathways become more pronounced as the zebrafish develops.

To explain the mechanisms behind the mitochondrial molecular changes observed in *xpnpep3^Δ7/Δ7^* zebrafish, we explored potential substrates of Xpnpep3. Previous studies in yeast and plants have identified several mitochondrial proteins processed by ICP55 and showed the importance of ICP55 in regulating mitochondrial protein quality [26, 27, 30]. Building on this knowledge, we compiled a comprehensive list of all known substrates of ICP55 and looked for their corresponding orthologues in zebrafish. We expanded this list by including zebrafish proteins that contained predicted ICP55/XPNPEP3 sites identified through computational analysis. Mathematical testing confirmed that these putative Xpnpep3 substrates were indeed found in reduced quantities in the mutant *xpnpep3^Δ7/Δ7^* larvae. Not surprisingly, these proteins play diverse roles within mitochondria, and many of them have been linked to mitochondrial disorders [58–60]. First, we mined our proteomics nLC-MS/MS data set for protein N-terminal semi-tryptic peptides and identified N-terminal peptides for several proteins of interest. Next, we determined which of these N-terminal peptides could be explained by misprocessing of the substrate under mutant conditions based on Xpnpep3’s predicted functional site. Our data revealed correct processing of Acadvl, Acat1, Glud1b, Pdha1a and Atp5f1b in wild-type replicates. Interestingly, in *xpnpep3^Δ7/Δ7^* larvae, we exclusively detected the non-cleaved peptide for Ndufab1b and Acadvl. All in all, these findings provide further evidence that the proposed substrates are actual targets of zebrafish Xpnpep3, supporting the conclusion that XPNPEP3 is the vertebrate orthologue of ICP55. We are particularly confident in Ndufab1b because: 1) Mitofates predicted it to have a Xpnpep3 cleavage site, 2) its yeast orthologue is a known substrate of ICP55, and 3) it is significantly downregulated in our analysis.

To validate that Ndufab1b and the aforementioned proteins are substrates of Xpnpep3, future research should incorporate techniques such as mitochondrial fraction enrichment and specialized mass spectrometry-based N-terminomics methods like COFRADIC, ChaFRADIC, or TAILS [61]. By fractionating and separately analyzing mitochondrial and cellular fractions, functional mitochondrial proteins can be distinguished from precursor proteins that are still in the cytosol undergoing synthesis or transport. Mass spectrometry-based N-terminomics selectively label and identify N-terminal peptides, enabling a more comprehensive detection of substrates by which we can more precisely pinpoint targets of Xpnpep3. In addition, it will be important to perform these studies in patient-derived samples from unrelated families.

Recently, also an Xpnpep3-deficient mouse model has been described [22]. The authors of this study focus on renal and ciliary functions and phenotypes because of the known link with NPHPL1. At 16 weeks of age, Xpnpep3-deficient mice displayed elongated primary cilia and disrupted mitochondrial morphology in the renal epithelial cells. Phenotypically, the mutant mice developed kidney tubular dilatation and fibrosis under stress conditions (i.e. cisplatin injection). Interestingly, the authors also reported reduced activity of respiratory complex I in mutant mice kidneys, along with decreased protein expression of various complex I subunits. These findings further support our results on the mechanisms and substrates of XPNPEP3 in maintaining the mitochondrial proteome integrity, including OXPHOS subunits, and overall mitochondrial health. This paper did not address the function or composition of other respiratory complexes, or explore additional tissues or phenotypes. It will be interesting to investigate whether these mice also display or develop other mitochondrial disease-related phenotypes, in particular cardiomyopathy.

Interestingly, mitochondrial intermediate peptidase (MIPEP), another intermediate peptidase involved in the processing of presequences to form mature mitochondrial functional proteins, is known to cause a mitochondrial disorder in humans (combined oxidative phosphorylation deficiency-31 (COXPD31); MIM 617228) [62–64]. As also outlined in the introduction, MIPEP removes octapeptides following MPP cleavage [65]. Studies using cells from patients with bi-allelic *MIPEP* variants have demonstrated impaired MIPEP function, leading to compromised processing of mitochondrial proteins, including various OXPHOS complex subunits and the mitochondrial insertase OXA1L [64], resulting in their decreased abundance and impaired overall mitochondrial function. Some patients’ skin fibroblasts showed respiratory chain dysfunction limited to complex I, others also had also issues with complexes III and IV [64]. Clinical features often seen include left ventricular noncompaction, global developmental delay, and hypotonia. Other variable symptoms can include seizures, cataracts, microcephaly, and cardiomyopathy. COXPD31 becomes apparent shortly after birth or in early infancy and can result in infantile death. Like in patients with bi-allelic *XPNPEP3* variants, cardiac involvement is variably present [62–64]. Thus, similar to *XPNPEP3*-related disease, the clinical and pathological characteristics among patients with COXPD31 are heterogeneous and can affect multiple organs. In contrast to COXPD31, which typically has an early onset, the onset of disease-related phenotypes in XPNPEP3 patients tends to occur later, with most reported cases presenting symptoms in late childhood or early adulthood [21–23]. Recently, an elderly man with a homozygous nonsense variant in *XPNPEP3* was reported, who presented with nephronophthisis and hearing loss but without cardiac involvement [23]. This suggests that LoF variants in *XPNPEP3* have a milder phenotype than bi-allelic *MIPEP* variants. This discrepancy in phenotypic severity and age of onset seems to be in accordance with the protein functions of MIPEP and XPNPEP3. The impaired removal of a larger number of destabilizing amino acids (octapeptides) by MIPEP is likely to cause greater protein instability and subsequent mitochondrial dysfunction, than the inefficient removal of a single destabilizing amino acid by XPNPEP3. Despite these insights, the underlying mechanisms causing these mechanistic and phenotypic differences in these diseases remain incompletely understood. The recognition of *XPNPEP3* as a disease-causing gene, and its inclusion to mitochondrial and cardiac disease gene panels will be essential for patient identification, to further understand the genotype-phenotype correlations and to gain better insights into the mechanisms driving this clinical variability.

In summary, we provide patient and zebrafish disease modeling evidence that XPNPEP3 is the vertebrate ortholog of ICP55, functioning in the processing and stabilization of mitochondrial precursor proteins and maintaining mitochondrial health. Omics analyses in the Xpnpep3-deficient zebrafish furthermore identified putative Xpnpep3 substrates, whose impaired processing might be at the molecular basis of the disease. Nonetheless, it will be important to further explore these findings in patient-derived samples from unrelated families. Finally, as our study identifies mitochondrial dysfunction as central to the disease, extended metabolomics studies in zebrafish and patient samples, could provide additional insights into the pathogenesis resulting from XPNPEP3 dysfunction. Specifically, these metabolic signatures can help with the diagnosis and management of the disease, and may aid in the identification of therapeutic targets.

## Methods

### Patients

#### Clinical assessment

The proband and first-degree family members underwent clinical assessment including a physical examination, 12-lead electrocardiogram, and transthoracic echocardiography. CMR in sibling III:1 was performed on a clinical 1.5T SIGNA Artist system. NT-proBNP and hs-cTnT were directly measured in fresh serum samples using standard procedures. Elevated NT-proBNP was defined as >15 pmol/L [66]. Elevated hs-cTnT was defined as >14 ng/L [67]. Serum levels of FGF-21 were determined in duplicate using a commercial ELISA kit (Millipore).

#### Trio exome sequencing (ES)

Peripheral blood samples from the proband, sibling III:1 and parents were used for genomic DNA (gDNA) extraction through standard procedures. The Agilent SureSelect Human All Exon V7 Enrichment Kit version 7 was employed for capturing exons and flanking splice junctions. Sequencing was conducted on an Illumina platform, and reads were aligned to the human reference genome GRCh37/hg19 using BWA (http://bio-bwa.sourceforge.net/). Variant calling was performed using the GATK haplotype caller (https://www.broadinstitute.org/gatk/). The identified variant underwent annotation and filtering via Alissa Interpret software (Agilent). Priority was given to *de novo*, recessive, and X-linked rare variants (minor allele frequency <0.1% in public databases). Sanger sequencing was employed for the verification of the identified variant and for testing other family members.

#### Linkage analysis with ES data

Linkage analysis was performed with the ES data of the available family members. Single-nucleotide variants (SNVs) were hard-filtered for depth >10 and genotype quality >80. Additionally, only one SNV per 30kbp window was retained [68], resulting in 25,831 usable SNVs. The filtered variants were then annotated with HapMap phase II the recombination map [69]. Parametric multipoint linkage analysis was run in MERLIN [70] with an autosomal recessive inheritance model. Disease allele frequency in the population was set to 1/100,000 and penetrance to 0.0001 (reference/reference), 0.0001 (alternative/reference) and 1.0 (alternative /alternative). The known first-cousin consanguinity between the parents was also encoded into the model. Following linkage analysis, genomic regions were considered for further screening if they had a logarithm of odds (LOD) score >0.

#### Fibroblast cell culture

The fibroblasts obtained from skin biopsy were cultured using standard procedures in an ISO-certified laboratory within the Clinical Genetics department. The culture medium used was Ham’s F10, containing 15% (v/v) fetal calf serum (FCS) and 1% (v/v) penicillin/streptomycin (PS).

#### Nonsense-mediated mRNA decay (NMD) assay

To inhibit nonsense-mediated mRNA decay, patients and age-matched control fibroblasts were cultured in the presence of 100 μg/mL CHX (Sigma-Aldrich) for 24 hours. To isolate RNA, the cells were cultured in

75 cm^2^ tissue culture flasks until they reached 70%–80% confluency. The RNeasy mini kit (Qiagen, Venlo, the Netherlands) was used for RNA isolation according to the manufacturer’s protocol. cDNA was generated with the iScript cDNA synthesis kit (Biorad). Reverse transcription PCR was performed following standard protocols with primers indicated in **Table 2**.

**Table 2.**
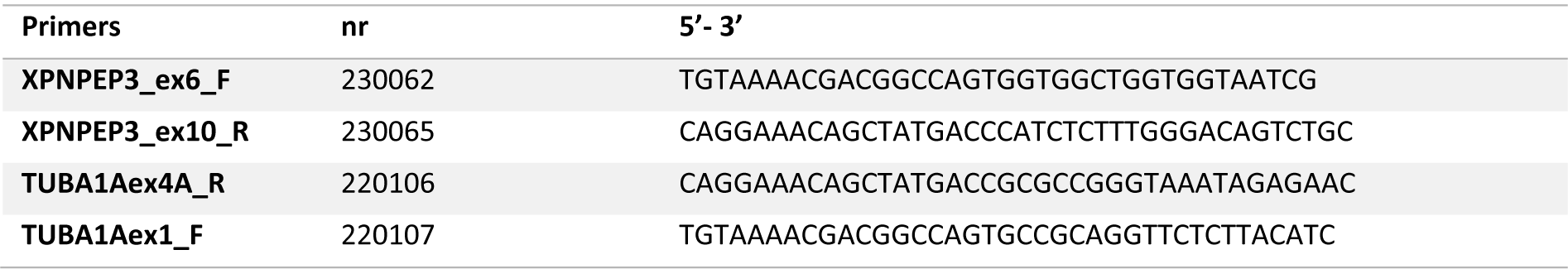
Primers used for RT-PCR analysis.

#### Western Blot analysis

Fibroblasts from affected siblings and age-matched controls were cultured in duplicate until reaching 70%–80% confluency. Protein extracts were prepared from fibroblast cultures as well as from frozen ventricular myocardial tissue of affected siblings using RIPA buffer supplemented with cOmplete Protease Inhibitor Cocktail (Roche). Protein concentrations in the lysates were determined using the Pierce BCA Protein Assay Kit. Five age-matched healthy control ventricular myocardial tissue lysates that were previously established were provided by Jolanda van der Velden (Amsterdam University Medical Center). Approximately 20 ug of total protein lysate was loaded on 4–15% Criterion TGX Stain-Free precast gel (Bio-Rad), and proteins were separated at 100V for 90 minutes. Thereafter, the gels were activated by exposure to UV light to visualize the total protein input **(Supplemental Figure 3)**. Proteins were then transferred to a nitrocellulose membrane using the Trans-Blot Turbo Transfer System (Bio-Rad). Membranes were blocked using 5% skim milk powder/TBST and incubated with 1:1500 primary antibody (α-XPNPEP3, 15655-1-AP, Proteintech) overnight at 4 °C. Thereafter, blots were washed twice with TBST for 5 minutes, 15 minutes with 0.5M NaCl, followed by two additional 5-minute washes in TBST. Secondary antibody (IRDye 800CW Goat anti-Rabbit) was added at a 1:10000 dilution. Washing steps were repeated as before, with the addition of a final 5-minute rinse in TBS. The membrane was imaged with an Odyssey CLX scanning system (Li-Cor).

#### Cardiac histology and ultrastructural analysis

Heart samples obtained at HTx from left and right ventricular wall and septum were formalin-fixed and paraffin-embedded using standard clinical laboratory protocols. For light microscopy, paraffin-embedded tissue was cut at 4 µm, mounted on charged slides and stained for hematoxylin and eosin (H&E) and Masson’s trichrome. For electron microscopy examination, myocardial tissue of the explanted tissue was fixed in 2% paraformaldehyde and 2.5% glutaraldehyde in 0.1M phosphate buffer (pH 7.4), embedded in Epon and cut in 70 nm sections. Images of the idiopathic DCM, NF donor and HCM myectomy samples were acquired as part of previous studies [53, 71].

#### Mitochondrial complex enzyme activity measurements

Measurements of the mitochondrial oxidative phosphorylation enzyme activities were performed in mitochondria-enriched fractions from patients’ fibroblasts and cardiac tissue according to established procedures [72]. For these experiments, fibroblasts from healthy controls and cardiac tissue from patients without underlying mitochondrial disease were used as controls.

### Zebrafish model

#### Zebrafish handling

Zebrafish (*Danio rerio*, strain: Tüb/AB) husbandry was conducted in accordance with institutional guidelines and national animal welfare legislation, following standard conditions. Adult wild-type zebrafish were kept under typical laboratory conditions as previously outlined. Zebrafish embryos were cultured at 28°C in egg water, composed of 1 M HEPES-buffered (pH 7.2) E3 medium (34.8 g NaCl, 1.6 g KCl, 5.8 g CaCl2 · 2H2O, 9.78 g MgCl2 · 6 H2O). To prevent pigmentation, 0.2 mM 1-phenyl-2-thio-urea (PTU, Thermo Fisher Scientific, Waltham, United States) was introduced to the egg water at 24 hours post-fertilization (hpf) for high-speed imaging.

#### Generation of xpnpep3 zebrafish mutant

The Alt-R™ CRISPR/Cas9 System RNA of IDT was used to create single-guide RNAs (sgRNAs) with target sequence 5’ CAGTTTTGCCCTCATCACGTGGG 3’. The Alt-R crispr RNA (crRNA) and Alt-R trans-activating crispr RNA (tracrRNA) were combined in a 1:1 ratio in Duplex Buffer for annealing. The mixture was subjected to incubation at 95 °C for 5 minutes, followed by cooling to room temperature. To form the ribonucleoprotein complex, 50 pmol of sgRNA was mixed with 4 ng of Cas9 protein and incubated for 5 minutes at room temperature. Approximately 1.0 nL was microinjected into the yolk of 1-cell–stage embryos. Injected embryos were raised to adulthood and outcrossed for germline transmission. To assess successful genome editing and germline transmission F1 embryos of F0 founder were screened for out-of-frame deletions. To do so, A DNA fragment covering the CRISPR target site was amplified by PCR and sequenced by Sanger sequencing using primers xpnpep3_in8Fw_c and xpnpep3_in9Rv **(Table 3)**. After mutant selection, allele-specific primers were designed and used for genotyping **(Table 3)**.

**Table 3.**
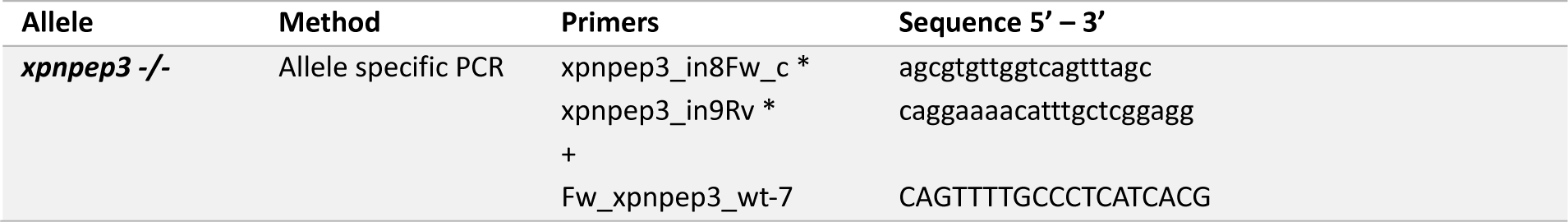
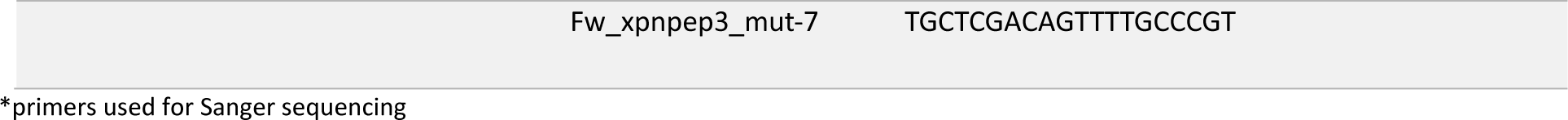
Genotyping details of *xpnpep3* mutant zebrafish.

#### Zebrafish genotyping

Adult zebrafish were subjected to anesthesia using 0.016% Tricaine, followed by excision of a small portion of the caudal fin. Larvae were anesthetized and harvested individually. Lysis was performed in 80 µl of 50 mM KOH. Genomic DNA from both zebrafish larvae and adult tissue was isolated by incubating at 95°C until complete tissue dissolution. Post-incubation, 8 µl of 1M Tris-HCl at pH 8 was added. The resulting samples were centrifuged, and 1 µl of the sample was employed for allele-specific PCR. For allele-specific PCR, 1.0 μL of 10 μM allele-specific primer was also added to the reaction mixture (in a 1:1:1 ratio with general primers **(Table 3)**. PCR conditions were used as previously described [38].

#### Touch response assay

Zebrafish larvae were placed individually into a well plate containing 20 ml E3. Touch response was provoked with a 23G needle. Videos were recorded with the Olympus SZX116 microscope (Olympus) with a DP72 camera (Olympus) and analysed afterwards. Response was categorized as, 1) normal: upon first touch larvae immediately start swimming or 2) delayed: if larvae required additional time to start swimming or if they required more than one provocations before swimming.

#### Cardiac analysis

Larvae were anesthetized using 0.02% Tricaine and mounted in a ventral position in 0.25% (w/v) agarose (Sigma-Aldrich) on glass-bottom Petri dishes. 10-second videos were captured at 100 frames/s using a high-speed camera (CrashCam™, IDT Europe) mounted on an Axio Observer inverted microscope (Zeiss) with Stage Top heating system (Ibidi) at 28°C. Cardiac volumes and contractility parameters were derived by manually outlining the perimeter of the ventricle at end-diastole and end-systole [38, 73, 74].

#### Mitochondrial complex enzyme activity measurements

At 5 dpf, *xpnpep3^Δ7/Δ7^* and wild-type larvae were collected. Per genotype, for each of the three biological replicates, 80 larvae were pooled together. Samples were snap-frozen in liquid nitrogen and stored at -80 °C until further use. Upon thawing, samples were homogenized in 1450 μL of 10mM Tris–HCl buffer at pH 7.6. Subsequently, 250 μL of 1.5 M sucrose was added, and the mixture was subjected to centrifugation at 14,000 g for 10 minutes at 2°C. The supernatant was discarded, and the resulting pellet was suspended in 200 μL of 10mM Tris-HCl buffer at pH 7.6. Enzymatic activities of complex I-IV, citrate synthase, and the protein content of the samples were assessed spectrophotometrically as previously described [40, 72].

#### RNA sequencing (RNA-Seq) and data analysis

At 5 dpf, *xpnpep3^Δ7/Δ7^*and wild-type larvae were collected. Per genotype, for each of the three biological replicates, 20 larvae were pooled together. Total RNA was then isolated from these triplicates using TRIzol™ Reagent (ThermoFisher Scientific). Total RNA was checked for quality on an Agilent Technologies 2100 Bioanalyzer using an RNA nano assay. All samples had RNA integrity number (RIN) values greater than 9.7. Triplicate RNA-seq libraries were prepared using the Illumina TruSeq Stranded mRNA protocol (www.illumina.com). Cluster generation was conducted following the Illumina TruSeq SR Rapid Cluster Kit v2 (cBot) Reagents Preparation Guide (www.illumina.com). Detailed descriptions for both procedures are described in [75]. After hybridization of the sequencing primer, sequencing-by-synthesis was performed using the NextSeq2000 with a paired end read 50-cycle protocol followed by dual index sequencing.

<u>Data analysis:</u> Cutadapt [76] was used to trim off Illumina adapter sequences from the reads, which were subsequently mapped against the GRCz11 zebrafish reference using HiSat2 (v2.2.1) [77]. Gene expression values were called using htseq-count (v0.12.4) [78] and Ensembl release 103 gene and transcript annotation. Differential expression analysis of count data was performed in Rstudio (v2024.04.0+735) [79] with the DEseq2 (v1.42.1) [80] and tidyverse package (v2.0.0) [81]. Differentially expressed genes (DEGs) between the *xpnpep3^Δ7/Δ7^* and wild-type conditions were identified based on the following criteria: a false discovery rate (FDR) of <0.05 and an absolute log2FC of > 1. Volcano and principal component analysis (PCA) plots were generated in R studio using the tidyverse package (v2.0.0) [81]. For readability, annotations were manually added in Adobe Illustrator v25.1. For detailed descriptions of advanced data analysis methods, such as GSEA, please refer to the Supplemental Materials, in section “**Advanced omics data analysis**”.

#### Mass spectrometry and data analysis

At 5 dpf, *xpnpep3^Δ7/Δ7^*and wild-type larvae were collected. Per genotype, for each of the three biological replicates, 5 larvae were pooled together, snap-frozen and snap-frozen in liquid nitrogen and stored at - 80 °C until further use. Upon thawing, triplicate samples were transferred to a fresh Eppendorf and 200 µL 50 mM ammonium bicarbonate (ABC) and 0.5 % (v/v) sodium deoxycholate (SDC) was added. The samples were sonicated for 30 min and incubated for 10 min at 85 °C. The samples were centrifuged for 10 min at 10,000 × g to pellet the eye pigment. Protein concentrations were measured using the BCA assay (Thermo Scientific). 100 µg protein was extracted by acetone precipitation for 60 min at -20 °C. Samples were centrifuged at 14,000 × g for 20 min, acetone was removed, and the pellet was allowed to air dry. The protein pellet was dissolved in 200 μL 50 mM ABC and 0.5 % (v/v) SDC for 30 min in a sonication bath. Proteins were digested with LysC (1:100 enzyme:protein ratio) twice for 3 hrs at 37 °C and 1100 rpm. Subsequently, 10 µL 50% immobilized trypsin slurry (Immobilized Trypsin, TPCK Treated, Thermo) was added and the digestion proceeded overnight at 30 °C and 1150 rpm. Digests were acidified with 0.5 % (v/v) final concentration trifluoroacetic acid (TFA) and centrifuged at 14,000 × g for 20 min to remove the precipitated SDC. The supernatant was transferred to a new tube. 10 µg of the digests were purified using an in-house packed StageTip with two Empore^TM^ Octadecyl C18, 47 mm disks (Supelco Analytical), dried using a SpeedVac Vacuum concentrator, and stored at -20 °C.

Nanoflow liquid chromatography-tandem mass spectrometry (nLC-MS/MS) was performed on an EASY-nLC coupled to an Orbitrap Eclipse Tribid mass spectrometer (Thermo Fisher Scientific), both operating in positive mode. The samples were suspended in 2 % (v/v) ACN, 0.5 % (v/v) FA to reach a final concentration of 0.4 µg/µL, based on the BCA assay. 800 ng per sample were injected onto and separated on an analytical column (75 μm × 35 cm ReproSil-C18 reversed-phase column, Dr Maisch, 2.4 μm, packed in-house) a 120 min segmented gradient from 3 % to 20 % buffer B (buffer A = 0.1 % (v/v) FA; buffer B = 80 % (v/v) ACN,

0.1 % (v/v) FA) for 70 min, 20 % to 35 % buffer B for 30 min at a flow rate of 350 nl/min using and 35% to 80% buffer B for 5 min with 500 nl/min The column was kept at 50 °C in a NanoLC oven -MPI design (MS Wil GmbH). Electrospray ionization was performed using a 1.8 kV spray voltage and a capillary temperature of 275 °C. The mass spectrometer was operated in data-dependent acquisition (DDA) mode. The spectra were acquired in the Orbitrap at 60K resolution for a maximum injection time of 118 ms with an automatic gain control (AGC) target of 400000. High-resolution HCD MS2 spectra were generated using a normalized collision energy of 30 % with 15K. The AGC target was set to 100 % and the maximum injection time to “Auto”.

<u>Data analysis:</u> The resulting raw files were analyzed using Spectronaut (SN, v. 18.6, Biognosys) with a directDIA+ search against the zebrafish UniProt database (UP000000437, release December 2022) using the following settings: Trypsin was set as the digestion enzyme, allowing for up to 2 missed cleavages. N-terminal protein acetylation and methionine oxidation were set as variable modifications (allowing up to 5 variable modifications per peptide). Mass tolerances were left as dynamic, and the precursor, peptide, and protein FDR were set to 0.01. The quantity MS-level was set to MS2, and data imputation was disabled. The protein LFQ method was set to MaxLFQ, and the major (Protein) and minor (Peptide) group quantities were determined by summed peptide and precursor quantities respectively. Protein group inference was performed using the IDPicker algorithm. A full overview of the settings is given in the Supplemental Materials, in section **“Detailed settings Spectronaut”**.

Protein quantities were normalized by way of median centering. Standard Student’s t-tests were performed on normalized log-transformed protein quantities. Proteins were considered suitable for differential gene expression analysis if five out of the six replicates (i.e. triplicates from *xpnpep3^Δ7/Δ7^*mutant and wild-type samples) presented protein quantities. Conversely, proteins that did not meet this criterion were determined unsuitable and were excluded from statistical analysis. Normalization and statistical analyses were performed in Visual studio code using Python v3.11 with the SciPy package (v1.13.0). Differentially expressed proteins (DEPs) between the *xpnpep3^Δ7/Δ7^*and wild-type conditions were identified based on the following criteria: a p-value of <0.05 and an absolute log2FC of > 1. Detailed descriptions of all advanced data analysis methods, such as GSEA, are provided in the Supplemental Materials in section “**Advanced omics data analysis**”.

#### Putative Xpnpep3 substrates

Putative Xpnep3 substrates were identified through a combination of literature review and computational analysis using the Mitofates prediction tool [43]. The literature review resulted in a list of proteins which included experimentally determined ICP55 substrates of Saccharomyces cerevisiae [26, 27, 42], substrates identified in Arabidopsis thaliana [30] and putative human XPNPEP3 substrates [20]. For each of these proteins, we manually searched for the best matching *Danio rerio* orthologue and identified it as a putative Xpnpep3 substrate. For the computational analysis using Mitofates, the full Uniprot Danio rerio reference proteome was downloaded (canonical and isoforms) and protein sequences were used as input. For every sequence input, this tool generated a score indicating the likelihood of containing a mitochondrial presequence and the presence of an ICP55 cleavage site. Proteins scoring above 0.5 for mitochondrial presequences and showing a predictable ICP55 cleavage site were considered as a putative Xpnpep3 substrate. In total, we ended up with a list of 166 putative substrates, of which 146 were suited for statistical analysis **(Online Supplemental Data file 5)**.

To determine whether the group of putative Xpnpep3 substrates were downregulated in the proteomic data set, a bootstrap analysis was performed. The mean log2FC of 100000 randomly selected protein sets (containing n=146 proteins, which contains the same number of proteins that were identified as putative Xpnpep3 substrates) was denoted. For each protein set a one-tailed Student’s t-test was performed between the putative Xpnpep3 substrate group and random protein set of that iteration. Hα= mean log2FC of Xpnpep3 substrate group < mean log2FC random protein set.

#### Search to detect peptides that are indicative of Xpnpep3’s enzymatic activity

To detect peptides that are indicative of Xpnpep3’s enzymatic activity, or indicate compromised activity in mutant conditions, a second directDIA+ search was conducted specifically to identify processed protein N-termini. A specialized database was created for this search by filtering the complete zebrafish UniProt database to focus exclusively on putative Xpnpep3 substrates, as identified by the Mitofates tool. This filtered database was then used to generate all semi-tryptic peptides with a tryptic C-terminus and a non-tryptic N-terminus, permitting up to 3 missed cleavages. Additionally, peptides were required to have a minimum length of 6 amino acids and be derived from the first 100 amino acids of the protein sequences. This approach resulted in a file containing protein-specific peptide quantities for all 6 replicates. Next, we filtered this dataset based for peptides whose N-termini were either localized to the Xpnpep3 cleavage site or had one amino acid extra upstream from that site. To collect high confidence substrates, this group of proteins was next filtered based on a probability score of containing mitochondrial presequences provided by Mitofates of ≥0.25. The fragmentation mass spectra of individual peptide sequence identifications from this group of proteins were manually reviewed in Spectronaut. Peptide identifications based on insufficient sequence coverage were excluded from further analysis. By comparing the peptide profiles from the six replicates, we could identify peptides that could be specifically explained by the activity (or lack thereof) of Xpnpep3.

#### Statistical analysis

All statistical tests regarding omics analyses were performed in R studio v2024.04.0+735 with R build 3.3.0. Statistical analysis regarding other zebrafish experiments were performed using GraphPad Prism v.6 software and consisted of the statistical test stated in the figure legends. Results are expressed as mean ± SD, unless otherwise indicated. For our study, a value of P <0.05 was considered statistically significant. Whenever possible, masking was performed in data collection and analysis.

## Data availability

All data produced in the present study are available upon reasonable request to the authors.

## Acknowledgements

We wish to thank the patients and their family members for participation in this study. We thank Anne Werkhoven and Rashida Ghazi from the Clinical Genetics Department of the Erasmus MC for their excellent assistance. We thank dr. Prateek Arora and prof. Nadia Mercader Huber at the University of Bern for kindly providing the basis of the zebrafish Mitocarta inventory. Prof. Jolanda van der Velden at the Amsterdam University Medical Center for providing the age-matched healthy control ventricular myocardial tissue lysates used for Western Blot analysis. Dr. Michiel Dalinghaus at the Erasmus MC and prof. Kenneth B. Margulies at the University of Pennsylvania for kindly sharing pediatric tissue samples and dr. Nicole N. van der Wel at the Amsterdam University Medical Center for providing assistance with electron microscopic analysis. Lastly, we would like to thank the animal caretakers at the Erasmus MC for zebrafish husbandry. This work was supported by the Dutch Heart Foundation (grant number 03-003-2020-T062 awarded to Judith M.A. Verhagen).

