## Supplemental Material for "Bi-allelic variants in the aminopeptidase XPNPEP3 cause mitochondrial disease with pediatric cardiomyopathy"

### Supplemental Figures

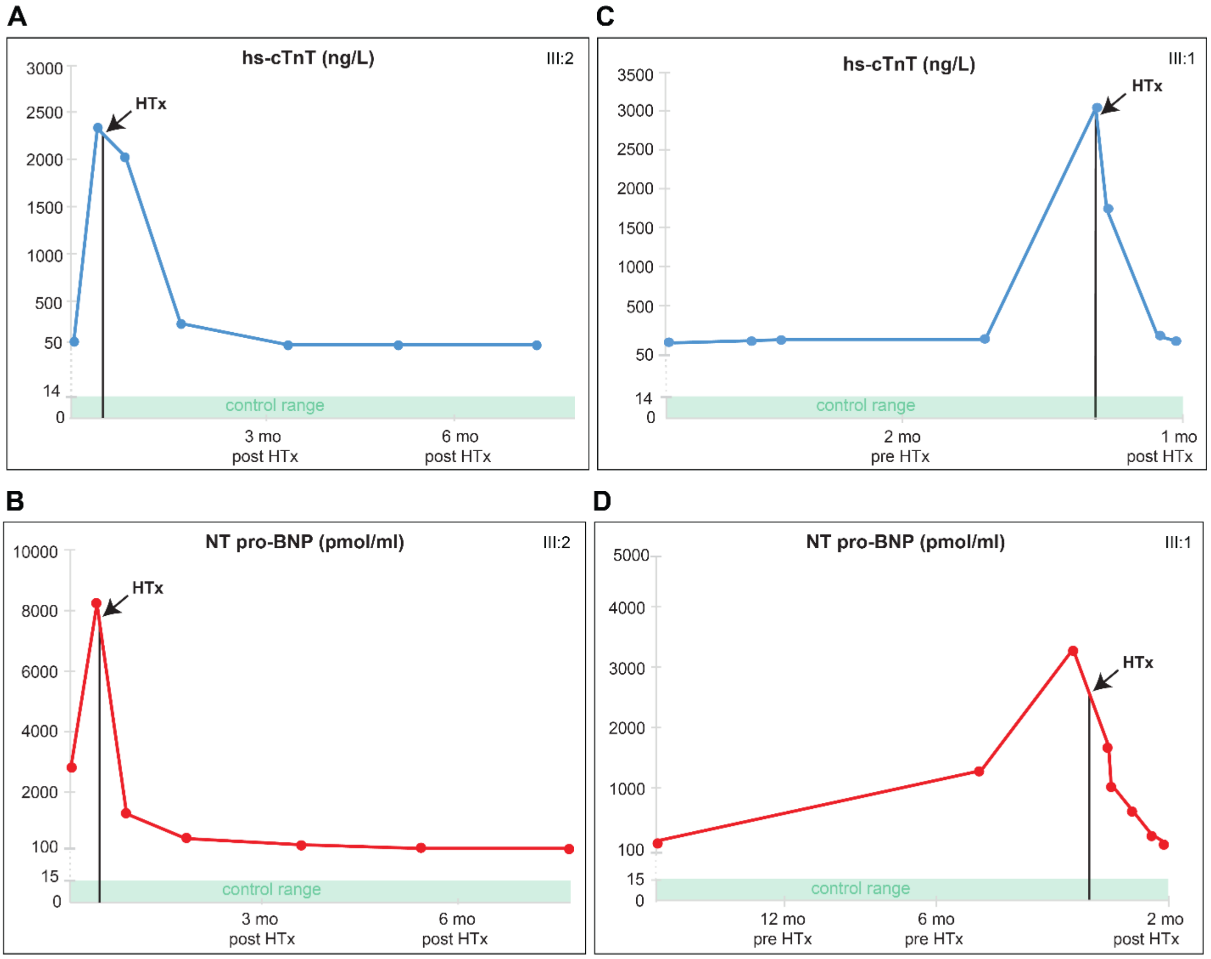


**Supplemental Figure 1. High-sensitivity cardiac troponin T (hs-cTnT) and N-terminal pro-B-type natriuretic peptide (NT-proBNP) levels in affected siblings.** (A-C) Levels were measured in blood before and after heart transplantation (HTx). hs-cTnT levels of the proband III:2 in (A). NT-proBNP levels of the proband III:2 in (B). hs-cTnT levels of sibling III:1 in (C). NT-proBNP levels of sibling III:1 in (D). Control ranges (<15 pmol/ml for NT-proBNP and <14 ng/L for hs-cTnT) are indicated in green.

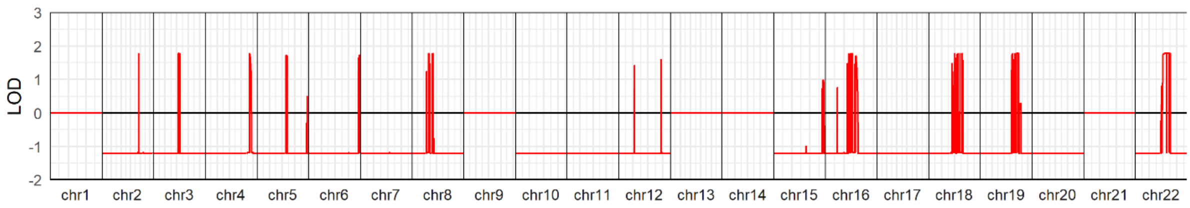


**Supplemental Figure 2. Linkage analysis.** The highest logarithm of the odds (LOD) detected is 1.7794 (not significant) and there are 120 regions supported by the linkage analysis. The variant c.1357G>A in *XPNPEP3,* located at position chr22:41320486G>A, marks one of the breakpoints of a linkage-supported region, with a score of 1.7794.

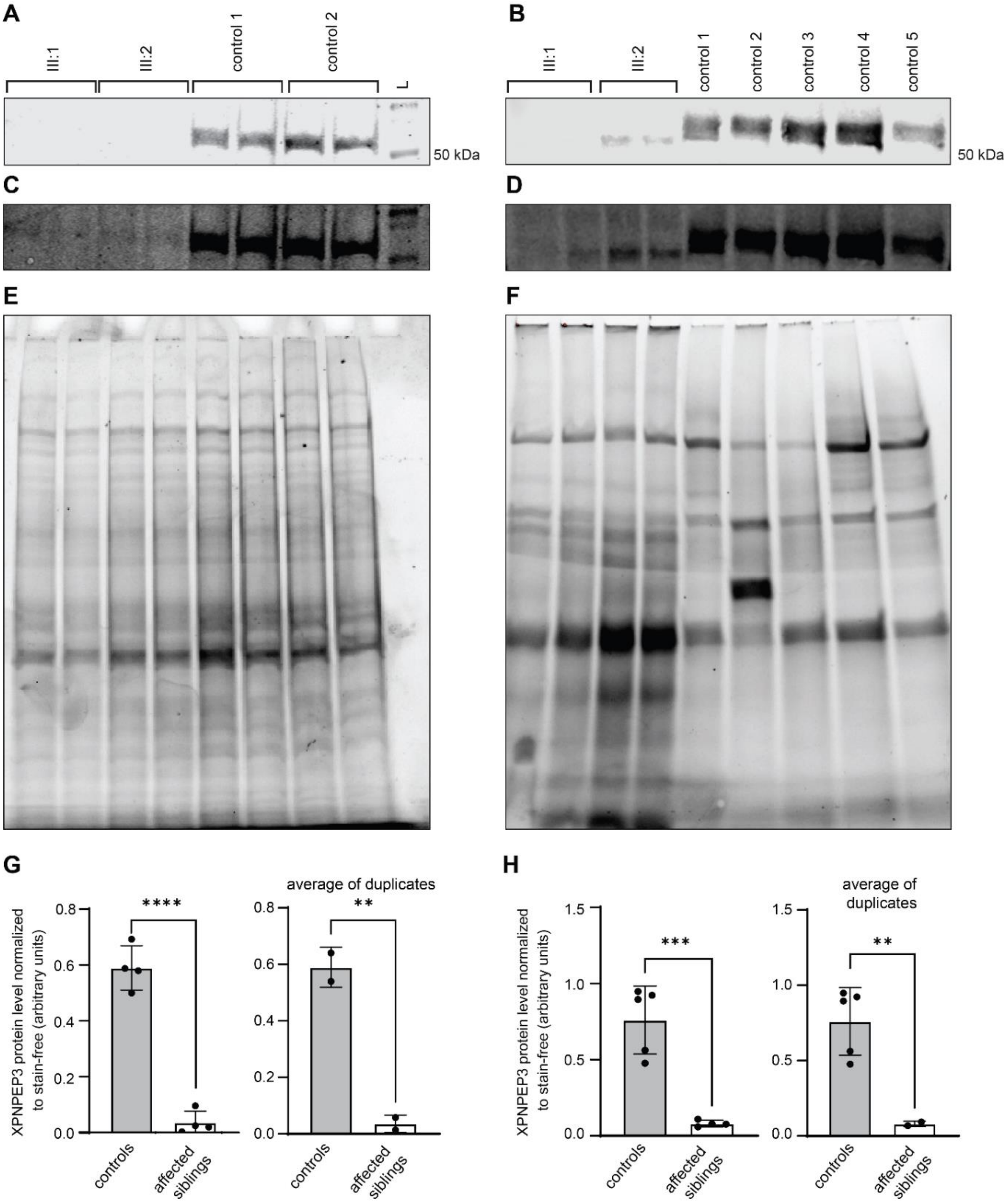


**Supplemental Figure 3. Quantification of XPNPEP3 expression levels normalized to Stain-Free input.** (A-B) Western blots showing XPNPEP3 expression in fibroblasts (A) and cardiac tissue (B). (C-D) Same blots as in A-B, respectively, but with adjusted brightness and contrast to enhance the visualization of any faint bands. (E-F). Total protein input visualized by UV illumination of Criterion TGX Stain-Free gels. The images in these panels correspond to the gels from which the Western blots in panels A-D were derived. (G-H) Bar plots representing the relative expression levels of XPNPEP3 across the different samples with error bars representing standard deviation. To ensure fair comparison among samples, XPNPEP3 protein levels were quantified and normalized to the total protein as visualized by the Criterion TGX Stain-Free image. The right graph displays the average of duplicates, where applicable. Statistical significance was determined using an unpaired Student's t-test, with $p < 0.05$ indicating significant differences.

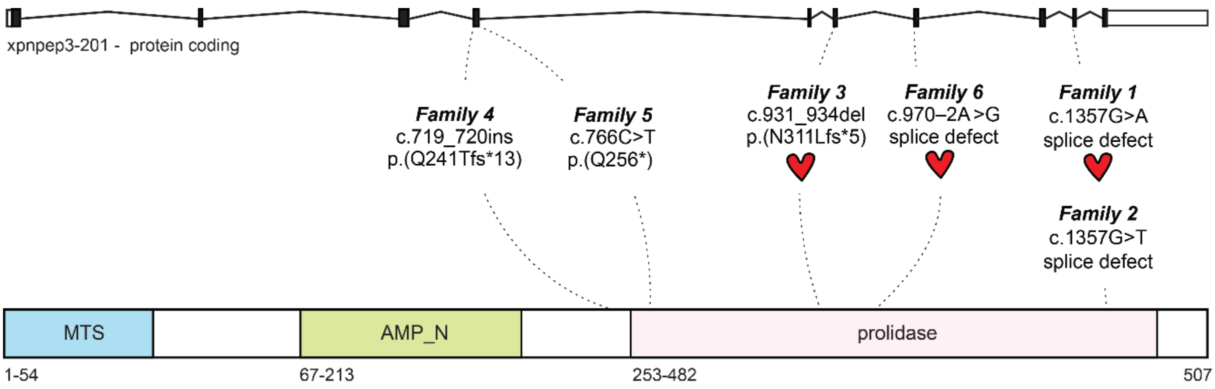


**Supplemental Figure 4. Schematic representation of the *XPNPEP3* gene, protein structure and location of reported homozygous loss-of-function variants**. The *XPNPEP3* gene contains 10 exons, that encode for the XPNPEP3 protein of 507 amino acids having a mitochondrial target signal (MTS), amino peptidase domain (AMP_N) and prolidase domain. Reported homozygous are shown. These variants are all located near or in the prolidase domain. Families with cardiomyopathy are represented by a heart symbol.

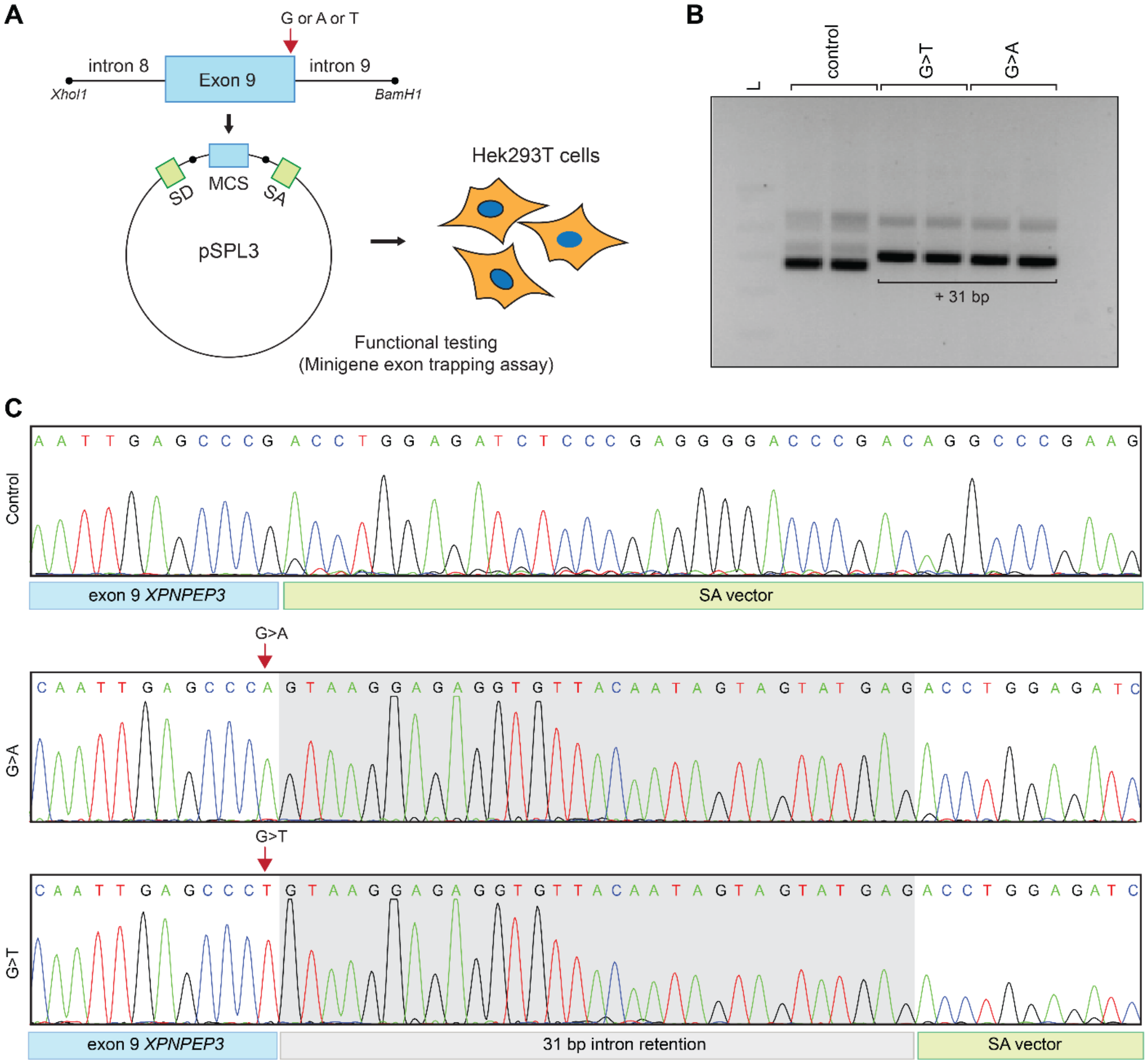


**Supplemental Figure 5. Splicing minigene trapping assay reveals identical pattern of variant-induced mis-splicing between c.1357G>A and c.1357G>T.** (A) Schematic representation of the minigene splicing assay based on the pSPL3 exon trapping vector. The pSPL3 vector contains a splice donor (SD) and splice acceptor (SA) site that operate as exons and a functional intron. Wild-type exon 9 of *XPNPEP3* with partial flanking introns, or site-mutagenesis derived mutant fragments with the c.1357G>A variant (family reported in this paper) or c.1357G>T variant (family A131 [1], referred to as family 2 in this paper), were separately cloned into the *XhoI* and *BamHI* sites of the vector. Indicated reporter minigenes were transfected in HEK cells. (B) Gel electrophoresis of RT-PCR products derived from the three reporter minigenes. (C) Sequencing trace depicting use of alternative splice donor site, leading to 31 bp intron retention.

**A** CLUSTAL O(1.2.4) multiple sequence alignment

```
homo            MPWLLSAPKLVPAV----ANVRGLSGCMLCSQRRY--SLQPVPERRIPNRYLGQPSPFTH 54
danio           --MFHSSSGLLRAAVQCASRSYWSPGSVWCPCRHVSVKTEGWNSKKVPQRYLGQPSPYTH 58
                   : *:  *: *.    :.     *.: *  *:   . :   .:::*:********:**

homo            PHLLRPGEVTPGLSQVEYALRRHKLMSLIQKEAQGQSG------TDQTVVVLSNPTYYMS 108
danio           PHLIRHGEVTPGLTQTEYELRRQRLASLIEIQAERQTGSGASSNSSHIVIILSHPIRYMS 118
                ***:* *******:*.** ***::* ***: :*: *:*      :.: *::**:*  ***

homo            NDIPYTFHQDNNFLYLCGFQEPDSILVLQSLPGKQLPSHKAILFVPRRDPSRELWDGPRS 168
danio           NDIPYPFHQNQDFLYLTGITEPDSALVMYGSS----KPDQAVLFVPRRDPARELWDGPRS 174
                ***** ***:::**** *: **** **: .        .:*:********:*********

homo            GTDGAIALTGVDEAYTLEEFQHLLPKMKAETNMVWYDWMRPSHAQLHSDYMQPLTEAKAK 228
danio           GKDGAAALTGLERVHSTEELGVVLKSIK--GGTVWYDNSQPCHPRLHQTHVRPLLEGGQ- 231
                *.*** ****::..:: **:  :* .:*   . ****  :*.* :**. :::** *.

homo            SKNKVRGVQQLIQRLRLIKSPAEIERMQIAGKLTSQAFIETMFTSKAPVEEAFLYAKFEF 288
danio           ---LVKSLRPLTHSLRAIKSPAEVALMKEAGRITAQAFKKTMAMSRGNIDEAVLYAKFDY 288
                    *:.:: * : ** ******:  *: **::*:*** :**  *:. ::**.*****::

homo            ECRARGADILAYPPVVAGGNRSNTLHYVKNNQLIKDGEMVLLDGGCESSCYVSDITRTWP 348
danio           ECRAHGANFLAYPPVVAGGNRANTLHYINNNQIVKDGEMVLLDGGCEYFGYVSDITRTWP 348
                ****:**::************:*****::***::*************   **********

homo            VNGRFTAPQAELYEAVLEIQRDCLALCFPGTSLENIYSMMLTLIGQKLKDLGIMKNI-KE 407
danio           VNGKFSAAQRELYEAVLEVQLACLSQCSPGVSLDYIYSTMLTLLARQLKELGILPSHASD 408
                ***:*:* * ********:*  **: * **.**: *** ****:.::**:***: .  .:

homo            NNAFKAARKYCPHHVGHYLGMDVHDTPDMPRSLPLQPGMVITIEPGIYIPEDDKDAPEKF 467
danio           TDAMKAARQFCPHHVGHYLGMDVHDTPELSRSQPLQPGMVITIEPGLYISEDNRSCPERF 468
                .:*:****::*****************:: ** *************:** **::..**:*

homo            RGLGVRIEDDVVVT-QDSPLILSADCPKEMNDIEQICSQAS- 507
danio           RGLGVRIEDDVVIRDHGGPLILSANTPKTISEVERTCAHAED 510
                ************:  :..******: ** :.::*: *::*.
```

**B**

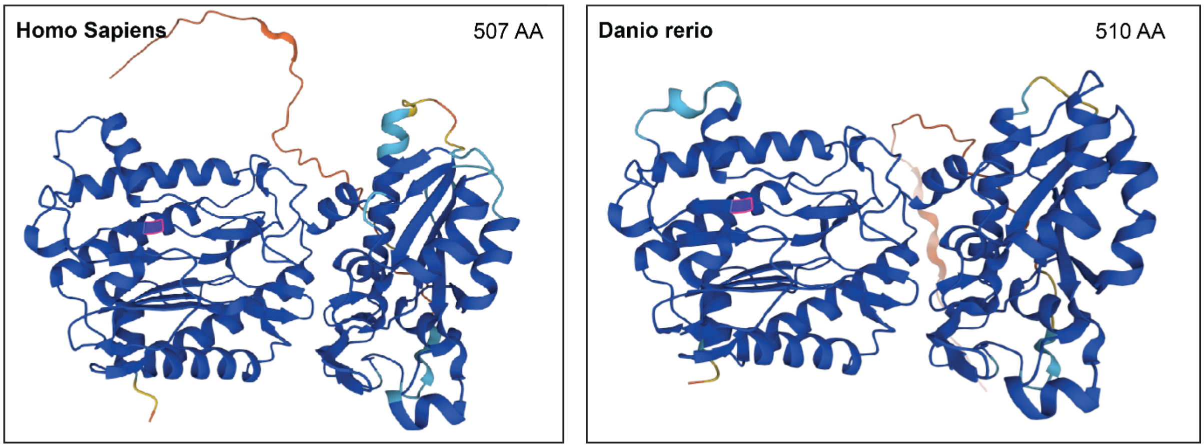


**Supplemental Figure 6. Comparative analysis of XPNPEP3 proteins in humans and zebrafish.** (A) Amino acid sequence alignment of XPNPEP3 from humans and zebrafish using Clustal Omega. (B) Tertiary structure of the human and zebrafish XPNPEP3 proteins as predicted by Alphafold-2 with accession numbers: Q9NQH7and A8KBZ9, respectively.

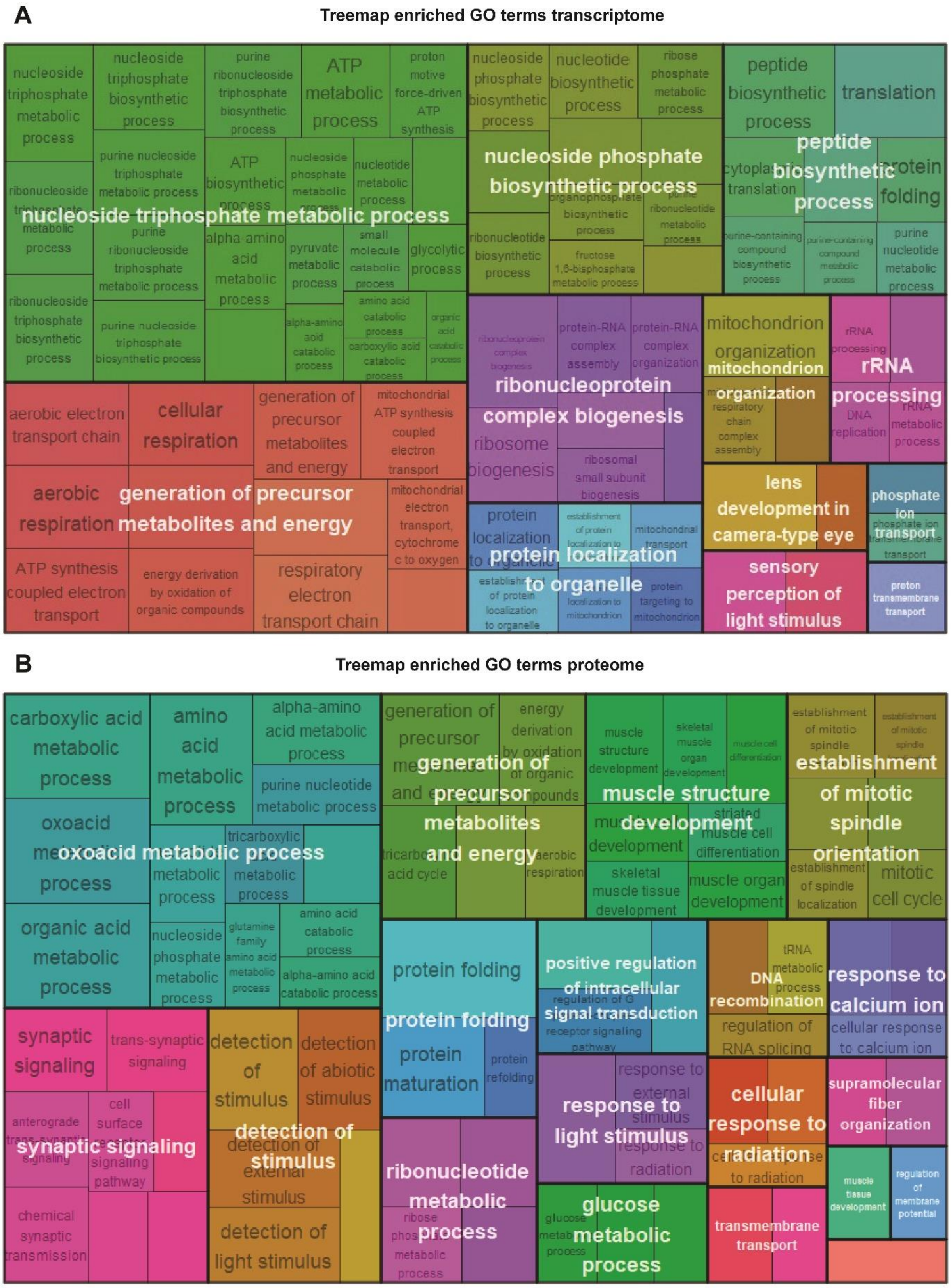


**Supplemental Figure 8. Treemap visualization of the GSEA-based enriched GO terms.** (A) Treemap visualization of the enriched GO terms in transcriptomic data, based on the GSEA. (B) Treemap visualization of the enriched GO terms in the proteomic data, based on the GSEA. Each annotated box in the treemap corresponds to a GO-enriched term. The size of each parent box indicates the significance of enrichment, with larger parent boxes representing higher -log10 adjusted p-values. The relative size of the child term boxes within the treemap reflects their significance relative to their parent term.

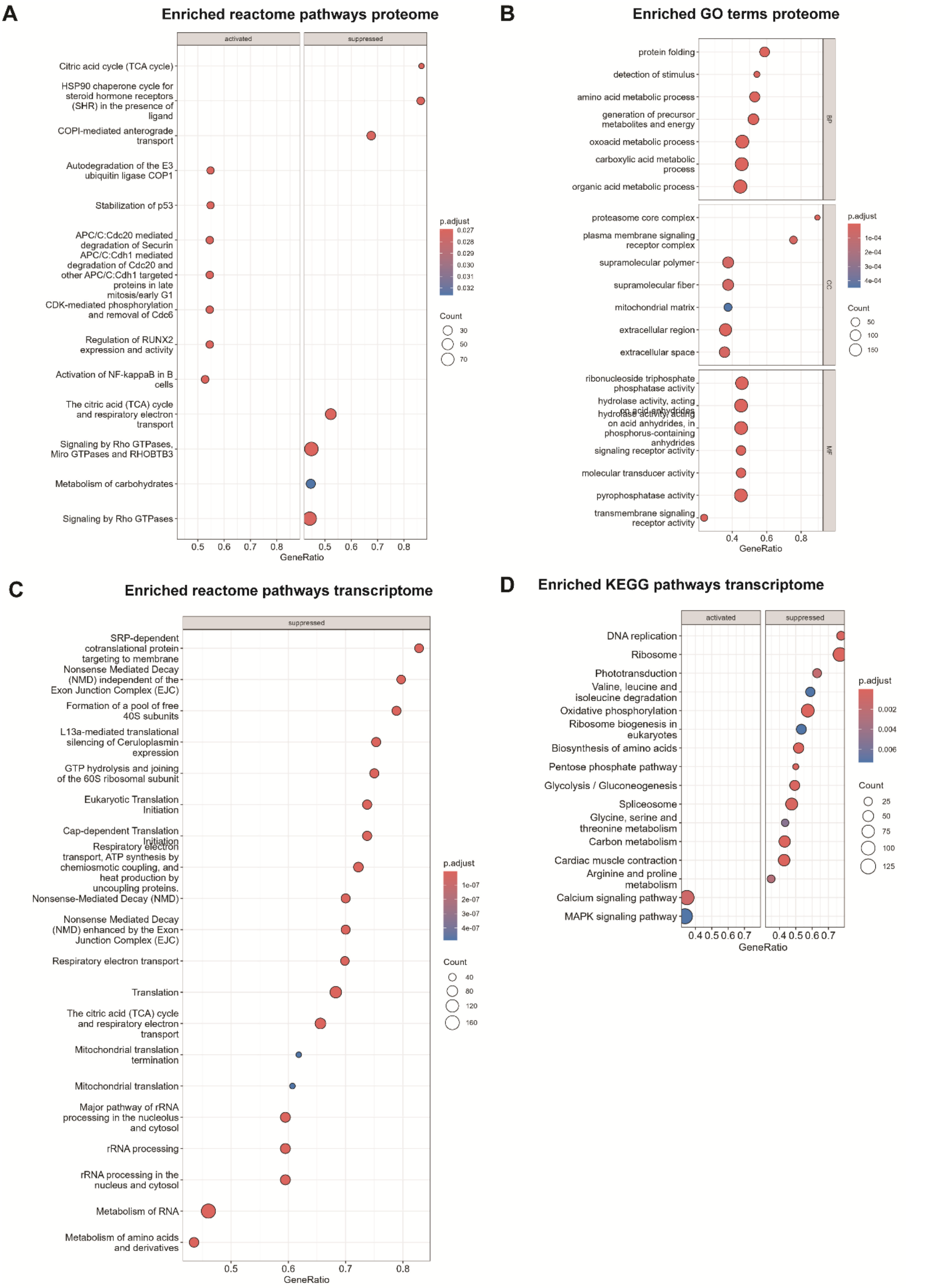


**Supplemental Figure 9. Gene Set Enrichment Analysis.** (A) Dot plot visualization of GSEA-derived enriched reactome pathways in proteomic data. (B) Dot plot visualization of the GSEA-derived GO terms in the proteomic data. The top term biological processes (BP), cellular compartment (CC) and molecular function (MF) are shown. (C) Bubble plot visualization of GSEA-derived enriched reactome pathways in transcriptomic data. (D) Dot plot visualization of GSEA-derived enriched KEGG pathways in transcriptome data. For all dot plots, the x-axis shows ratio of genes to number of genes in the term. The size of each circle denotes the counts of genes in the term and colour denotes adjusted p-value.

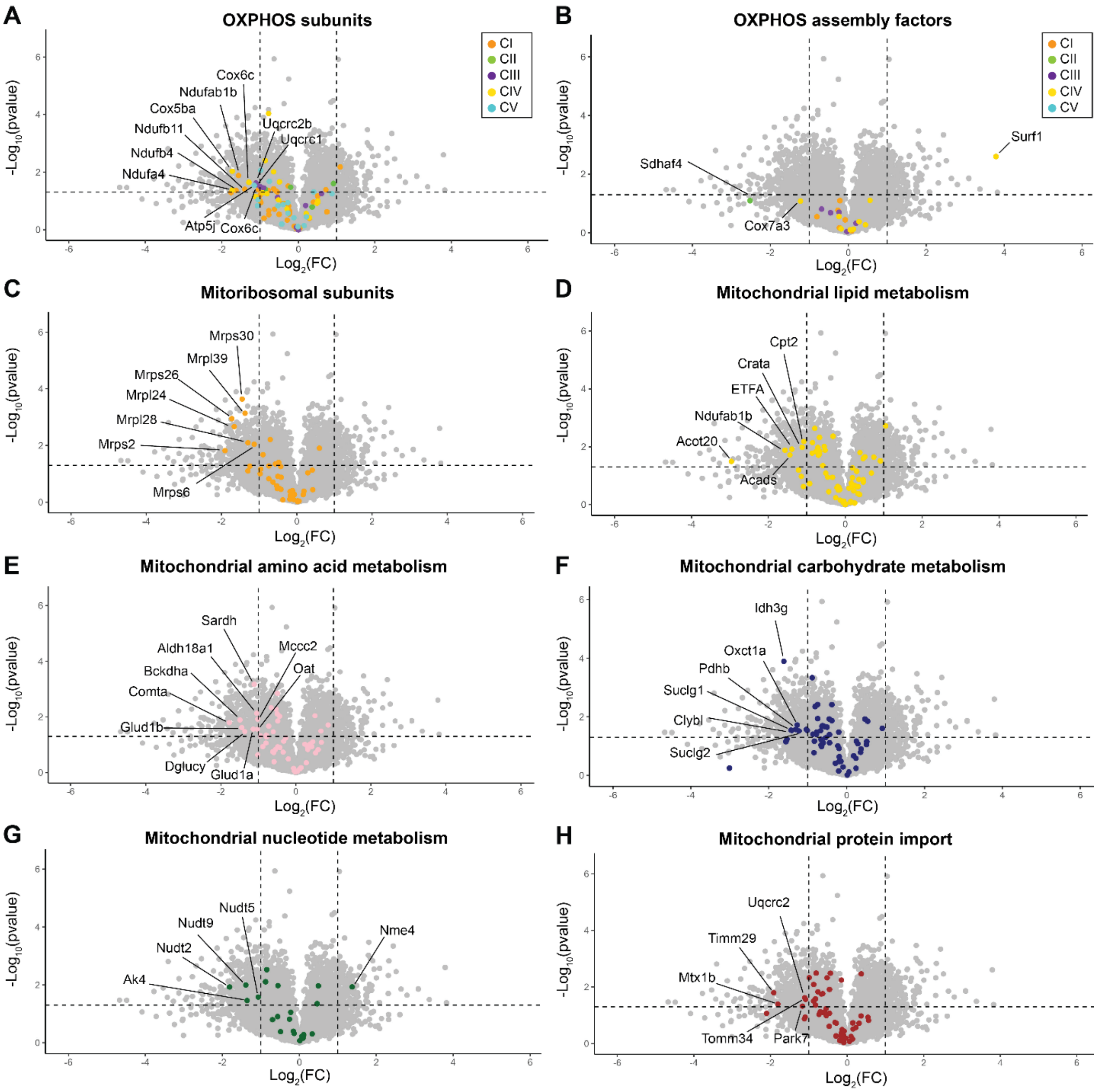


**Supplemental Figure 10. Volcano plots comparing mitochondrial proteins quantified in *xpnpep3*$^{\Delta7/\Delta7}$ zebrafish mutants relative to wild-types at 5 dpf.** Mitochondrial proteins were categorized according to the MitoCarta list and coloured as indicated. Subunits of OXPHOS complexes are shown in (A), OXPHOS assembly factors are shown in (B), mitoribosomal subunits in (C), mitochondrial proteins involved in mitochondrial lipid metabolism in (D), mitochondrial proteins involved in amino acid metabolisms in (E), mitochondrial proteins involved in carbohydrate metabolism in (F), mitochondrial proteins involved in nucleotide metabolism in (G) and proteins involved in mitochondrial protein import are depicted in (H). Mitochondrial proteins showing differential expression, determined by a p-value <0.05 and an absolute Log2FC > 1.0-fold, were labelled.

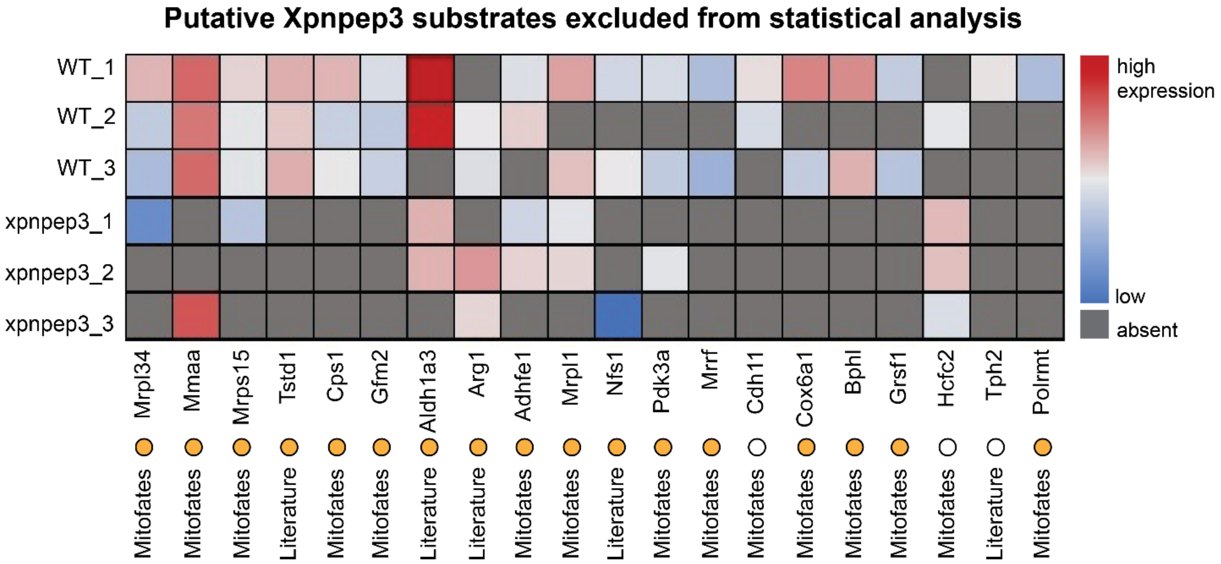


**Supplemental Figure 11. Putative Xpnpep3 substrates excluded from differential expression analysis due to insufficient data.** Among 166 putative substrates, 20 proteins lacked protein quantities in ≥5 replicates and were excluded from statistical analysis. Yet, the low or reduced expression of these proteins under mutant conditions may be attributed to Xpnpep3 dysfunction. Proteins of particular interest are those expressed in 3/3 wild-types and 0/3 xpnpep3$^{\Delta7/\Delta7}$ mutants, and include: thiosulfate sulfurtransferase like domain containing 1 (Tstd1), carbamoyl phosphate synthetase (Cps1) and GTP dependent ribosome recycling factor mitochondrial 2 (Gfm2). The yellow circle denotes mitochondrial proteins (based on the generated MitoCarta inventory).

### Supplemental Tables

**Supplemental Table 1. Clinical characteristics**

| Individual | Age at presentation (age range) | Presenting symptom | LVEF at presentation | Age at HTx | LVEF pre-HTx | CKD stage | Renal ultrasound |
|---|---|---|---|---|---|---|---|
| **III:1** | 11-15 | Asymptomatic | 44% | Within two years after presentation | 27% | CKD2 | Normal |
| **III:2** | 11-15 | Dyspnoea, exercise intolerance | 17% | Within four weeks after presentation | 11% | CKD3b | normal |

HTx, heart transplantation; eGFR, estimated glomerular filtration rate; CKD, chronic kidney disease (+stages).

**Supplemental Table 2. Molecular findings of trio-whole exome sequencing in the proband (III:2)**

| Gene | Accession | Nucleotide | Amino acid | Zygosity | MAF gnomAD v4.1.0 | CADD | AMCG Class | Evidence |
|---|---|---|---|---|---|---|---|---|
| *XPNPEP3* | NM_022098.4 | c.1357G>A | p.(Gly453Serfs18*) | Hom | 0.00058 | 36 | LP | PVS1, PM2_sup |
| *TTN* * | NM_001267550.1 | c.74225G>A | p.(Arg24742His) | Het, *de novo* | 0.00001 | 23.4 | VUS | PM2_sup PP2 BP4 |
| *DMD* * | NM_004006.3 | c.10086+3A>G | p.? | X-linked, hemi † | Absent | Absent | VUS | PM2_sup |

*absent in III:1; † heterozygous in II:2

Abbreviations: Hom, homozygous; het, heterozygous; hemi, hemizygous; LP, likely pathogenic; VUS, variant of unknown significance

**Supplemental Table 3. Reported bi-allelic *XPNPEP3* variants in 12 individuals from 8 unrelated families**

| Fam | ID | Nucleotide | RNA | Amino acid | Zygosity | AMCG | Evidence | Renal | Cardiac | Other | Ref |
|---|---|---|---|---|---|---|---|---|---|---|---|
| 1 | III:1 | c.1357G>A | r.1357_1358ins1357+1_1357+31 | Splice alteration and frameshift p.(Gly453Serfs*18) | Hom | P | PVS1, PM2_sup | CKD2 | HCM/ DCM | B-thalassemia, microcytic anaemia, hyperparathyroidism, tremor upper extremities | This paper |
| | III:2 | c.1357G>A | r.1357_1358ins1357+1_1357+31 | p.(Gly453Serfs*18) | Hom | P | PVS1 PM2_sup | CKD3b | HCM/ DCM | | This paper |
| 2 | A131 (II:1) | c.1357G>T | r.1357_1358ins1357+1_1357+31 | Splice alteration and frameshift p.(Gly453Cysfs*18) | Hom | P | PVS1, PM2_sup | CKD3b, cysts | HTN | Essential tremor | [1] |
| | A131 (II:3) | c.1357G>T | r.1357_1358ins1357+1_1357+31 | p.(Gly453Cysfs*18) | Hom | P | PVS1, PM2_sup | CKD3b, NPHP-like, cysts | HTN | Essential tremor, arachnoid cyst, sensorineural hearing loss | [1] |
| | A131 (II:4) | c.1357G>T | r.1357_1358ins1357+1_1357+31 | p.(Gly453Cysfs*18) | Hom | P | PVS1, PM2_sup | CKD3b, NPHP-like | HTN | Essential tremor, sensorineural hearing loss, muscle fatigue | [1] |
| 3 | F543 (II:1) | c.931_934del | - | p.(Asn311Leufs*5) | Hom | P | PVS1, PM2_sup | CKD5, NPHP-like | HCM/ DCM | Seizures, mental and developmental delay | [1] |
| | F543 (II:2) | c.931_934del | - | p.(Asn311Leufs*5) | Hom | P | PVS1, PM2_sup | CKD5, NPHP-like | HCM/ DCM | Seizures, mental and developmental delay | [1] |
| 4 | II:2 | c.719_720insA | - | p.(Gln241Tfs*13) | Hom | P | PVS1, PM2_sup | CKD | NR (age range: 11-15 years) | Anaemia, growth failure | [2] |
| 5 | | c.766C>T | - | p.(Gln256*) | Hom | P | PVS1, PM2_sup | CKD5 | No (age range: 51-55 years) | | [3] |
| 6 | | c.970–2A>G | c..970_1055del | Splice alteration p.(Asp324Valfs*10) | Hom | P | PVS1, PM2_sup | CKD | HCM/ DCM | Intellectual disability, epilepsy | [4] |
| 7 | (A-II-1) | c.634G>A † | - | p.(Ala212Thr) | Comp Het | VUS | PM2_sup, BP4 | NPHP-like | No (age <1 year) | None | [5] |
| | | c.761G>T † | - | p.(Arg254Leu) | | VUS | BS1, BP4, BP6 | | | | |
| 8 | (B-II-1) | c.-87C>T † | - | upstream | Comp Het | VUS | PM2_sup, BP7 | NPHP-like, renal cysts, proteinuria, hematuria | No (age range: 11-15 years) | Bone cysts | [5] |
| | | c.1261C>G† | - | p.(His421Asp) | | VUS | PM2_sup, PP3 | | | | |

† These variants, unlike the reported variants in the other families, are not associated with loss-of-function effects

Fam, family; Hom, homozygous; comp het, compound heterozygous; LP, likely pathogenic; NR, not reported; P, pathogenic; CKD, chronic kidney disease (+stage); DCM, dilated cardiomyopathy; HCM, hypertrophic cardiomyopathy; NPHP-like; nephronophthisis-like nephropathy

### Supplemental Methods

#### Mini gene splicing assay: site directed mutagenesis and minigene trapping assay

To investigate the impact of a particular intervention on splicing, minigene exon trapping assays by amplifying the exon of interest and surrounding intronic sequences from genomic DNA was performed. The amplified fragments were subjected to site-directed mutagenesis and cloned into the pSPL3 vector (Thermo Fisher Scientific, Invitrogen) via Gibson assembly. All constructs were verified by Sanger sequencing. Subsequently, 1 µg of the minigene construct was transfected into HEK 293T cells, which were maintained at 70%-90% confluency in a 12-well plate and transfected with 5 µg of polyethyleneimine. Following 24 hours of incubation, RNA isolation was carried out, and cDNA was synthesized using the iScript cDNA synthesis kit (BioRad). The products transcribed from the vector were amplified using a set of standard primers, and the RT-PCR products were analyzed by ethidium bromide gel electrophoresis. Primer sequences used in the minigene exon trapping assay and site-directed mutagenesis are provided in **Supplemental Table 5**.

**Supplemental Table 5. Primers used for mini-gene exon trapping and SDM**

| | |
|---|---|
| **Gibson cloning** | |
| Gibson-M13F-XPNPEP3 | tgtaaaacgacggccagtGGAGGGATTAAACTGAAGTGGC |
| Gibson-M13R-XPNPEP3 | caggaaacagctatgaccGAGAATTGCTGAACCTGGGG |
| **SDM** | |
| SDM-R g121a | tgtaacacctctccttactgggctcaattgtgattac |
| SDM-F g121a | Gtaatcacaattgagccc**a**gtaaggagaggtgttaca * |
| SDM-R g121t | tgtaacacctctccttacagggctcaattgtgattac |
| SDM-F g121a | Gtaatcacaattgagccc**t**gtaaggagaggtgttaca * |
| **Control primers cloning** | |
| pSPL3-MCS_F | CACTTGTGGAGATGGGGGTG |
| pSPL3-MCS_R | TTCTTGTGGGTTGGGGTCTG |
| **RT-PCR** | |
| pSPL3-SD6-fwd_F | CTGAGTCACCTGGACAACC |
| pSPL3-SA2-rev_R | ATCTCAGTGGTATTTGTGAGC |

* nucleotides highlighted in red are targeted for SDM

#### Advanced omics data analysis

1. Mapping

In order to be able to perform downstream GSEA analysis, the UniprotKB IDs needed to be mapped to both NCBI (Entrez) and Ensembl IDs, because UniprotKB IDs cannot be used as input for these analyses. First, we used the Uniprot ID mapping website tool [6], which successfully mapped 5002 UniprotKB IDs to Ensemble IDs and 11010 to NCBI IDs. For those UniprotKB IDs that only mapped to one of the two databases, and additional conversion step using the org.Dr.eg.db package (v3.18.0) was used [7]. This allowed us to convert NCBI IDs to Ensembl IDs and vice versa. As a result, we obtained 11074 UniprotKB IDs with a successfully mapped NCBI ID and 11028 UniprotKB IDs with a successfully mapped Ensembl ID **(Supplemental Figure 12)**.

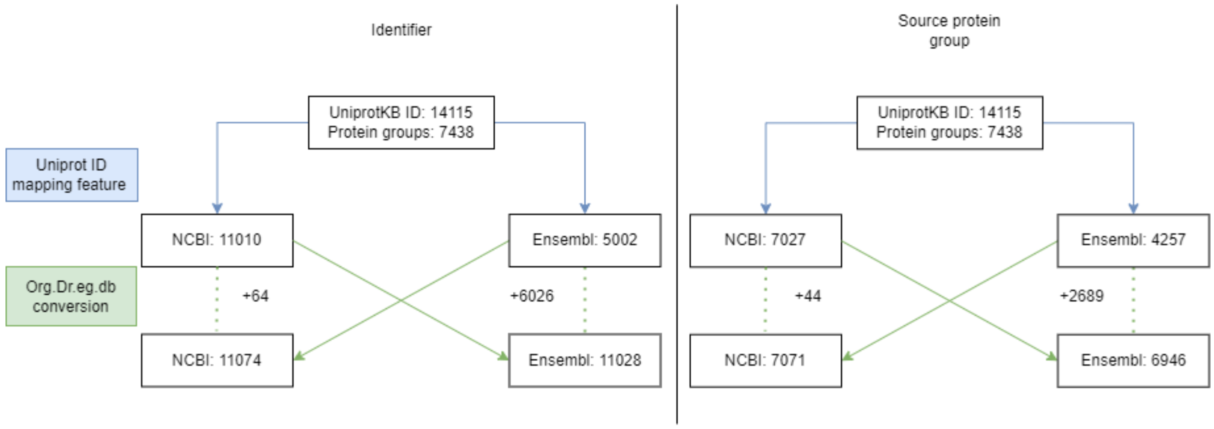


**Supplemental Figure 12. Strategy used to map UniprotKB IDs to NCBI and Ensembl IDs needed for downstream data analysis.** On the right also the number of protein groups corresponding to the number of mapped IDs is indicated.

Since each protein group could encompass one or more UniProtKB entries (proteins that were grouped together in one protein group as they shared the identified peptides), also one or more Ensembl ID could be mapped to a single protein group. For downstream analysis, one ID needed to be selected, which was initialized at random. However, as this project integrates proteomics and transcriptomic data, we refined the ID selection and chose the most appropriate ID when applicable. Specifically, if IDs from protein groups were present in the transcriptomics dataset, the random selection was restricted to these IDs.

To specify, out of the 7438 protein groups, 5439 contained only one Ensembl ID, which was used for further data processing. Notably, 5342 of these 5439 single-ID protein groups were also present as transcript IDs in the transcriptomics dataset. Additionally, 1507 protein groups contained multiple Ensembl IDs. Among these, 1306 protein groups had one Ensembl ID that matched a transcript ID in the transcriptomics dataset, and this ID was selected as the most appropriate ID. Out of the remaining 201 groups, 40 contained multiple Ensembl IDs, none of which matched any transcript IDs in the transcriptomic dataset. The remaining 161 groups contained multiple Ensembl IDs, of which 2 or more were present in the transcriptomic dataset and corresponded to different genes. For both of these groups, we randomly selected one Ensembl ID for further analysis. There were 492 protein groups that had no corresponding Ensembl IDs, and could not be used for further analysis.

2. Investigation of specific genes/proteins

To identify coordinated enrichments of gene sets, we ranked the transcriptomics dataset based on the test statistics (the Generalized linear model provided by DEseq2) and performed Gene Set Enrichment Analysis (GSEA). To identify a coordinated enrichments of protein sets, we ranked the proteomics dataset based on the test statistics (Students' t-test statistics for proteomics). The clusterProfiler package (v4.10.1) [8] was used to score enrichment of annotated sets in the GO database and Kyoto KEGG database. Ensembl IDs were used as input for GO-based GSEA and NCBI IDs were used for KEGG-based GSEA. The ReactomePA package (v1.46.0) was used to score enrichment of annotated sets in the Reactome database [9] using NCBI IDs. The Org.Dr.eg.db package (v3.18.0) was used for annotation of gene identifiers. The enrichPlot package (v1.22.0) [10] was used to generate dotplots and the rrvgo package (v1.14.2) [11] was used to generate the treemaps of the enriched GO terms, the algorithm of which is described in the rrvgo package documentation.

3. Mitochondrial genes/proteins based on the MitoCarta3.0 Inventory

The Mitocarta Inventory for *Danio rerio* was used to study the expression of the mitochondrial proteome. This inventory was based on the Human MitoCarta 3.0 inventory [12], and was kindly provided by Prateek Arora from the University of Bern. We updated the list by adding corresponding NCBI and Ensembl IDs to the zebrafish gene names, using our transcriptomic dataset as reference **(See Online Supplemental Data file 4)**. The differential expression of specific subsets of mitochondrial genes, based on the functional pathway annotations provided in the list, was visualized using Volcano plots generated in R studio using the tidyverse package (v2.0.0). Based on the inventory, all proteins encoding for OXPHOS and mitochondrial ribosomal subunits were selected, and their relative abundance was plotted. To calculate the relative complex abundances (RCA) of these subunits, normalized non-log2 transformed values were used. Normalization was done by dividing each replicate quantity by the respective replicate median. RCA were derived as the log2-transformed ratio of normalized *xpnpep3*$^{\Delta7/\Delta7}$ protein quantities to normalized wild-type protein quantities. The RCA plot was generated using the tidyverse package (v2.0.0).

**Detailed settings Spectronaut**

```
Settings Used:
 ├─ DIA Analysis\Calibration
 │  ├─ MZ Extraction Strategy:   Maximum Intensity
 │  ├─ Allow source specific iRT Calibration: True
 │  ├─ Precision iRT:      True
 │  │  ├─ Exclude De-amidated Peptides:  True
 │  │  └─ iRT <-> RT Regression Type:   Local (Non-Linear) Regression
 │  ├─ MS1 Mass Tolerance Strategy:     System Default
 │  └─ MS2 Mass Tolerance Strategy:     System Default
 ├─ DIA Analysis\Identification
 │  ├─ Precursor Qvalue Cutoff:  0.01
 │  ├─ Precursor PEP Cutoff:     0.01
 │  ├─ Protein Qvalue Cutoff (Experiment):    0.01
 │  ├─ Protein Qvalue Cutoff (Run):     0.01
 │  ├─ Protein PEP Cutoff:0.01
 │  ├─ Single Hit Definition:    By Stripped Sequence
 │  ├─ Exclude Single Hit Proteins:     False
 │  ├─ Exclude Duplicate Assays: True
 │  ├─ Exclude Predicted Fragment Scores:     False
 │  ├─ Generate Decoys:    True
 │  │  ├─ Decoy Generation Method:      Mutated
 │  │  │  └─ Preferred Fragment Source:   NN Predicted Fragments
 │  │  └─ Decoy Limit Strategy:   Dynamic
 │  │     └─ Library Size Fraction:      0.1
 │  └─ Pvalue Estimator:  Kernel Density Estimator
 ├─ DIA Analysis\Pipeline Mode
 │  ├─ Export All XICs:    False
 │  ├─ Generate SNE File:  True
 │  │  └─ Store Ion traces in SNE:      True
 │  ├─ Post Analysis Reports:
 │  │  ├─ CV Density Line Chart:  False
 │  │  ├─ CVs Below X Bar Chart:  False
 │  │  ├─ Data Completeness Bar Chart:   False
 │  │  ├─ Run Identifications Bar Chart:False
 │  │  └─ Scoring Histograms:     False
 │  │
```

```
│ ├─ PTM Report Schema:
│ ├─ Report Schema:     EXP2PEP (Normal), Peptide_Normal (Normal), Peptide_Pivot
(Pivot), Protein_Normal (Normal), Protein_Pivot (Pivot), BGS Factory Report (Normal)
│ └─ Reporting Unit:    Across Experiment
├─ DIA Analysis\Post Analysis
│ ├─ Differential Abundance Testing: Unpaired t-test
│ │ ├─ Assume Equal Variance:  False
│ │ ├─ Group-Wise Testing Correction: False
│ │ ├─ Log2 Ratio Candidate Filter:  0.58
│ │ └─ Confidence Candidate Filter:  Qvalue
│ │   └─ Confidence:    0.05
│ ├─ Differential Abundance Grouping: Major Group (Quantification Settings)
│ │ └─ Smallest Quantitative Unit:   Minor Group (Quantification Settings)
│ │   └─ Use All MS-Level Quantities: True
│ ├─ Calculate Explained TIC:  None
│ ├─ Calculate Sample Correlation Matrix:   False
│ └─ Hierarchical Clustering:  True
│   ├─ Distance Metric: Manhattan Distance
│   ├─ Linkage Strategy: Ward's Method
│   ├─ Order Runs by Clustering:     True
│   └─ Z-score Transformation: True
├─ DIA Analysis\Protein Inference
│ └─ Protein Inference Workflow:     Automatic
│   └─ Inference Algorithm:    IDPicker
├─ DIA Analysis\PTM Workflow
│ └─ PTM Localization:  True
│   ├─ Probability Cutoff:     0.75
│   └─ PTM Analysis:    True
│     ├─ Hierarchical Clustering:     True
│     ├─ Multiplicity:   True
│     ├─ Flanking Region: 7
│     └─ PTM Consolidation:     Sum
├─ DIA Analysis\Quantification
│ ├─ Precursor Filtering:      Identified (Qvalue)
│ │ └─ Imputation Strategy:    None
│ ├─ Proteotypicity Filter:    None
│ ├─ Protein LFQ Method: MaxLFQ
│ ├─ Quantity MS Level: MS2
│ ├─ Quantity Type:     Area
│ ├─ Cross-Run Normalization:  False
│ ├─ Quantification window:    Not Synchronized (SN 17)
│ ├─ Interference Correction:  True
│ │ ├─ Only Identified Peptides:     True
│ │ ├─ Exclude All Multi-Channel Interferences:   True
│ │ ├─ MS1 Min:   2
│ │ └─ MS2 Min:   3
│ ├─ Major (Protein) Grouping: by Protein Group Id
│ ├─ Minor (Peptide) Grouping: by Modified Sequence
│ ├─ Major Group Quantity:     Sum peptide quantity
│ ├─ Major Group Top N: False
│ ├─ Minor Group Quantity:     Sum precursor quantity
│ └─ Minor Group Top N: False
├─ DIA Analysis\Workflow
│ ├─ Method Evaluation: False
│ ├─ MS2 DeMultiplexing: Automatic
│ ├─ Multi-Channel Workflow Definition:     From Library Annotation
│ │ └─ Fallback Option: Labeled
│ ├─ Profiling Strategy: None
│ ├─ Run Limit for directDIA Library: -1
│ └─ Unify Peptide Peaks Strategy:   None
├─ DIA Analysis\XIC Extraction
│ ├─ XIC IM Extraction Window: Dynamic
│ │ └─ Correction Factor:     1
```

```
│ ├─ XIC RT Extraction Window: Dynamic
│ │ └─ Correction Factor:     3
│ ├─ MS1 Mass Tolerance Strategy:    Dynamic
│ │ └─ Correction Factor:     1
│ └─ MS2 Mass Tolerance Strategy:    Dynamic
│   └─ Correction Factor:     1
├─ Pulsar Search\Identification
│ ├─ PSM FDR:    0.01
│ ├─ Peptide FDR: 0.01
│ ├─ Protein Group FDR: 0.01
│ ├─ directDIA Workflow: directDIA+ (Deep)
│ └─ PTM Localization Filter:  True
│   └─ Min Localization Threshold:   0.75
├─ Pulsar Search\Labeling
│ └─ Channels:
│   ├─ Channel 1: False
│   ├─ Channel 2: False
│   └─ Channel 3: False
├─ Pulsar Search\Modifications
│ ├─ Max Variable Modifications:     5
│ └─ Select Modifications:
│   ├─ Fixed Modifications::
│   └─ Variable Modifications: :     Acetyl (Protein N-term), Oxidation (M)
├─ Pulsar Search\Peptides
│ ├─ Enzymes / Cleavage Rules: Trypsin/P
│ ├─ Digest Type: Specific
│ ├─ Max Peptide Length: 52
│ ├─ Min Peptide Length: 7
│ ├─ Missed Cleavages:  3
│ └─ Toggle N-terminal M:     True
├─ Pulsar Search\Result Filters
│ ├─ Fragment Ions:
│ │ ├─ Ion AA Length:   True
│ │ │ └─ N: 3
│ │ ├─ Ion Charge:      False
│ │ ├─ Ion Loss Type:   False
│ │ ├─ Ion Type:  False
│ │ ├─ m/z :      True
│ │ │ ├─ Max:     3000
│ │ │ └─ Min:     200
│ │ └─ Relative Intensity:     True
│ │   └─ Min:     1
│ └─ Precursors:
│   ├─ Amino Acids:     False
│   ├─ Best N Fragments per Peptide:  True
│   │ ├─ Max:     6
│   │ └─ Min:     3
│   ├─ Best N Peptides per Protein Group:   False
│   ├─ Channel Count:   False
│   ├─ FASTA Matched:   False
│   ├─ Missed Cleavage: False
│   ├─ Modifications:   None
│   ├─ Peptide Charge:  False
│   └─ Proteotypicity:  False
├─ Pulsar Search\Speed-Up
│ ├─ IM DFD Processing:
│ │ └─ Use Dynamic IM Peak Filter:   True
│ │   └─ Target TIC Fraction:  0.9
│ └─ MS2 Index:  Automatic
├─ Pulsar Search\Tolerances
│ └─ Tolerance Parameters:
│   ├─ Thermo IonTrap:
│   │ ├─ Calibration Search:   Dynamic
```

```
│   │ │ ├─ MS1 Correction Factor:    1
│   │ │ └─ MS2 Correction Factor:    1
│   │ └─ Main Search:   Dynamic
│   │   ├─ MS1 Correction Factor:    1
│   │   └─ MS2 Correction Factor:    1
│   ├─ Thermo Orbitrap:
│   │ ├─ Calibration Search:   Dynamic
│   │ │ ├─ MS1 Correction Factor:    1
│   │ │ └─ MS2 Correction Factor:    1
│   │ └─ Main Search:   Dynamic
│   │   ├─ MS1 Correction Factor:    1
│   │   └─ MS2 Correction Factor:    1
│   └─ TOF:
│     ├─ Calibration Search:    Dynamic
│     │ ├─ MS1 Correction Factor:     1
│     │ └─ MS2 Correction Factor:     1
│     └─ Main Search:    Dynamic
│       ├─ MS1 Correction Factor:     1
│       └─ MS2 Correction Factor:     1
└─ Pulsar Search\Workflow
   ├─ Fragment Ion Selection Strategy: Intensity Based
   ├─ In-Silico Generate Missing Channels:    False
   └─ Use DNN Predicted Ion Mobility:
```

### Online Supplemental Table Information

**Online Supplemental Data file 1.** Differential gene expression analysis of 5 dpf *xpnpep3*$^{\Delta7/\Delta7}$ larvae versus wild-types using DESeq2.

**Online Supplemental Data file 2.** Differential protein expression analysis of 5 dpf *xpnpep3*$^{\Delta7/\Delta7}$ larvae versus wild-types from mass spectrometry data.

**Online Supplemental Data file 3.** GSEA results for transcriptome and proteome data using GO, KEGG and Reactome databases.

- Sheet 1: GO transcriptome
- Sheet 2: KEGG transcriptome
- Sheet 3: Reactome transcriptome
- Sheet 4: GO proteome
- Sheet 5: KEGG proteome
- Sheet 6: Reactome proteome

**Online Supplemental Data file 4.** Mitocarta zebrafish inventory including differential expression values.

- Sheet 1: List of genes from the Human Mitocarta inventory with corresponding zebrafish genes, accession numbers, mitochondrial localization, and functional annotation
- Sheet 2: Mitocoarta genes present in transcriptomic data with differential expression values
- Sheet 3: Mitocarta proteins present in proteomics data with differential expression values

**Online Supplemental Data file 5**. List of putative Xpnpep3 substrates, corresponding differential protein expression analysis values, source of identification, and functional annotation

**Online Supplemental Data file 6**. Peptide-based analysis of putative substrates of Xpnpep3
